# Temporal trajectories of COVID-19 symptoms in adults with 22 months follow-up in a prospective cohort study in Norway

**DOI:** 10.1101/2024.04.30.24306604

**Authors:** Merete Ellingjord-Dale, Anders Nygaard, Nathalie C Støer, Ragnhild Bø, Nils Inge Landrø, Sonja Hjellegjerde Brunvoll, Mette Istre, Karl Trygve Kalleberg, John Arne Dahl, Linda Geng, Kostas Tsilidis, Elio Riboli, Giske Ursin, Arne Vasli Lund Søraas

## Abstract

**Objectives:** We aimed to describe the trajectories of cognitive and physical symptoms before, during, and after a positive- or negative SARS-CoV-2 test and in untested controls.

**Design:** A prospective cohort study.

**Setting:** Norway, 27 March 2020 to 6 July 2022

**Participants:** A total of 146 065 volunteers were recruited. Of these, 120 605 participants (mean age 49 (SD 13.7), 69% female), were initially untested for the SARS-CoV-2 virus, completed one or more follow-up questionnaires (response rates 72-90%) and were included for analysis. After 22 months of follow-up, 15 737 participants had a positive SARS-CoV-2 test, 67 305 a negative test, and 37 563 were still untested.

**Main outcome measures:** We assessed reported symptoms the past three weeks of memory or concentration problems, anosmia and dysgeusia, dyspnoea, fatigue, fever, headache, cough, muscular pain, nasal symptoms, sore throat and abdominal pain at baseline and through four follow-up questionnaires. In addition, overall health compared to a year before was measured with a five-point scale and memory problems were measured using the Everyday Memory Questionnaire-13 at two timepoints.

The exposure, SARS-CoV-2 test status (positive, negative or untested), was obtained from a mandatory national registry or from self-report, and data were analysed using mixed model logistic regression.

**Results:** A positive SARS-CoV-2-test was associated with the following persistent symptoms, compared with participants with a negative test (1-3 months after a negative test); memory problems (3 to 6 months after a positive test: adjusted odds ratio (OR) 9.1, 95% confidence interval (CI) 7.5 to 10.9; 12 to 18 months: OR 7.8, CI 5.7 to 10.8), concentration problems (3 to 6 months: OR 6.1, CI 4.8 to 6.5; 12 to 18 months: OR 5.3, CI 3.9 to 7.1), anosmia and dysgeusia, dyspnoea and fatigue as well as self-assessed worsening of overall health.

**Conclusion:** A positive SARS-CoV-2 test was associated with new onset memory- and concentration problems, anosmia and dysgeusia, dyspnoea and fatigue as well as self-assessed worsening of overall health, which persisted for the length of the follow-up of 22 months, even when correcting for symptoms before COVID-19 and compared to symptoms in negative controls.

**Trial registration:** ClinicalTrials ID: NCT04320732

**What is already known of this topic:**

- Cognitive difficulties and physical symptoms have been reported after infections with the SARS-CoV-2 virus, but lack of studies with data before and after infections have sparked a global debate regarding severity and duration of these symptoms.
- Post acute viral syndromes have been described after many different infections, but it is unknown whether the symptoms of “long-COVID” or Post Acute Sequela after COVID-19 (PASC) are specific to the SARS-CoV-2 or not.

**What this study adds:**

- We found a significant and long-term increase in cognitive symptoms, anosmia and dysgeusia, dyspnoea, fatigue, and self-assessed worsening of overall health after a positive (COVID-19) but not a negative (indication of another infection) SARS-CoV-2 test.
- To our knowledge, this is the first large cohort study to report the trajectories of both cognitive and physical symptoms from before and up to 22 months after a positive SARS-CoV-2 test, compared to SARS-CoV-2 negative controls.

## Introduction

The World Health Organization (WHO) has reported more than 770 million COVID-19 cases, including around 6.9 million deaths globally, as of August 25, 2023.^1^ It has been estimated that more than 65 million individuals around the world suffer from post-acute sequelae of severe acute respiratory syndrome coronavirus 2 (SARS-CoV-2) infection, often termed “long-COVID”.^2^ The definition of the phases of COVID-19 disease varies across studies.^3–8^ An acute phase 0-1 month after diagnosis, followed by a post-acute phase 1-3 months, and a post-COVID-19 phase >3 months have been described.^4^ The WHÒs description of the time frame for long-COVID coincides with the post-COVID-19 phase.^7^

Previous studies have reported a range of potential long-COVID symptoms such as anosmia and dysgeusia, fatigue, headache, muscle pain, abdominal pain, cough, dyspnoea, hair loss, and cognitive symptoms.^2,9–23^ However, most of these studies lacked relevant control groups, data on symptoms before COVID-19, and a long follow-up time. We initiated a large prospective cohort study in which we examined the temporal trajectories of symptoms in SARS-CoV-2-positive, - negative, and -untested participants, with a high response rate and a follow-up time of up to 22 months after the test date.

## Methods

### Study population

The Norwegian COVID-19 Cohort Study is a population-based, open cohort study of adult participants (aged 18-96 years) from Norway. From March 27, 2020, participants were recruited through social media, invitations, and nationwide media coverage. Participants completed a baseline questionnaire at inclusion and were invited to regular follow-up questionnaires. All participants had a Norwegian identification number and electronic access to the secure national digital governmental identification service.

Between March 27, 2020, to April 15, 2021, a total of 146 065 participants completed the baseline questionnaire (99 % before June 30, 2020). Of these, 127 798 participants were untested at baseline according to the mandatory Surveillance System for Communicable Diseases (MSIS) registry and confirmed this through self-report. We excluded 7193 participants who did not complete any follow-up questionnaires.

Our final study population of 120 605 had completed a baseline questionnaire and at least one follow-up questionnaire before July 6, 2022. The response rates for the first (May 2020), second (July 2020), third (November 2020), and fourth follow-up (December 2021) questionnaires were 79.3%, 84.2%, 80.4%, and 72.1%, respectively (Supplementary eFigure 1).

In a substudy between the 3^rd^ and 4^th^ follow-ups (July 2021), 6279 participants (all SARS-CoV-2-positive participants, and randomly selected negative and untested participants) were invited for an extra questionnaire (89.9% responded) (Supplementary eMethods 1). To assess the impact of non-response on our overall study, a random subset of these (n=2090) received telephone reminders if unresponsive electronically (“phone cohort”), with a 97% response rate in that group.

In another add-on study, from October 2020 to December 2021, 998 blood samples from 966 unvaccinated participants were collected and analyzed for SARS-CoV-2 antibodies (Supplementary eMethods 2).

The study was approved by the Norwegian Regional Committee for Research Ethics (REK 124170), and all participants submitted electronic informed consent forms. The study is registered in ClinicalTrials (https://clinicaltrials.gov; ID: NCT04320732), and reported according to the STROBE guidelines for cohort studies.

### Assessment of exposure

The exposure, SARS-CoV-2 status, was obtained through linkage of the participant’s personal identification numbers with the MSIS registry or self-report. COVID-19 became a compulsory reportable disease to MSIS on January 31, 2020. SARS-CoV-2 tests were done by a nasopharyngeal and/or oropharyngeal swab test and detected using real-time polymerase chain reaction (rt-PCR) in any accredited Norwegian clinical microbiology laboratory. From early in January, 2022, self-tests became increasingly available and PCR tests no longer mandated. SARS-CoV-2 status were therefore increasingly based on self-report the in the last months of the study. Participants could change their exposure status during the study period. We considered a participant untested from inclusion until the date of a SARS-CoV-2 test (positive or negative). If the first test was negative, a participant would be considered negative until the date of a later positive test. A participant with only negative tests was considered SARS-CoV-2-negative throughout the follow-up.

Community testing for SARS-CoV-2 status was free of charge and after the initial few months of the pandemic widely available and strongly encouraged. Testing for international travel was done at private facilities and required a fee and results were reported to MSIS like all other tests. Testing criteria changed during the study period and is described in Supplementary eFigure 2. From January 24, 2022, a PCR test was no longer routinely offered after a positive self-test.

### Assessment and definition of endpoints

Outcome measures were self-reported symptoms assessed through electronic follow-up questionnaires (Supplementary eMethods 3). Questionnaires were designed using existing knowledge about COVID-19 symptoms and the International Severe Acute Respiratory and Emerging Infection Consortium forms.^24^ For each questionnaire, participants were asked to check off symptoms they had experienced the past 3 weeks.

In the second follow-up questionnaire, a health transition question (“Compared to a year ago, or before you had COVID-19, how will you describe your health?”) with a 5-point Likert scale (the variable was dichotomized in the analyses), from the 36-Item Short Form Survey was added.^25^ As several participants reported memory problems in the free-text fields in the second follow-up questionnaire, questions on memory- and concentration were added to the third and fourth follow-up questionnaires and in the substudy between these questionnaires. To assess memory- and concentration problems, the Behaviour Rating Inventory of Executive Functions-adult version (BRIEF-A)^26^ was included in the substudy, while the 13-item Everyday Memory Questionnaire-revised version (EMQ-R)^27^ was included in the substudy and fourth follow-up.

### Statistical analyses

To investigate the temporal trajectory of symptoms after COVID-19, we examined the combined effect of SARS-CoV-2 status (untested, negative, or positive) and time since a SARS-CoV-2 test (0-1, 1-3,3-6, 6-12, 12-18 and >18 months) on symptoms. Time since test was defined as the time since the first positive or negative test, or the time since baseline (for the untested). We included information from all questionnaires, and used mixed-effect logistic regression calculating odds ratios (ORs) with 95% confidence intervals (CIs) of each symptom associated with SARS-CoV-2 status and time since test (eMethods 4). We compared symptoms of SARS-CoV-2-positive and untested participants with SARS-CoV-2-negative participants at one fixed time point, 1-3 months after their first negative test. We also compared the symptoms of SARS-CoV-2-positive and untested participants at each time point with the SARS-CoV-2-negative participants.

Analyses were adjusted for potential confounders: age (10-year categories), gender (men, women), body mass index (BMI,<25 kg/ m^2^ >25kg/ m^2^, missing), annual household income level (< 299 999, 300 000-599 999, 600 000-100 0000, >1 000 000 NOK, missing), smoking status (never, former, current, missing), underlying medical condition (no, yes, missing), and symptom status for each symptom at baseline (no, yes, missing) or at the first questionnaire asking about the symptom, but before the outcome symptom.

To illustrate the trajectories of symptoms before, during, and after a positive or negative test, we calculated the unadjusted moving average (with 95% CI) prevalence of symptoms for each exposure group (using the SARS-CoV-2-status at the last follow-up) over the whole study period (supplementary eMethods 5).

We performed the following sensitivity analyses and only including; participants with pre-omicron-variants, those who completed all questionnaires (not including the substudy), non-hospitalized participants, and participants with self-reported SARS-CoV-2 status.

In the substudy and fourth follow-up, we used multivariate analysis of variance to test for differences in subscales of BRIEF-A by SARS-CoV-2 status, and analysis of variance (ANCOVA) to test for differences by SARS-CoV-2 status on EMQ-R, controlling for age and sex. Pairwise-comparisons between SARS-CoV-2 status (untested, negative and positive) were Bonferroni corrected. We used ANCOVA to investigate the difference in change in EMQ-R between those who converted to positive and those who remained negative/untested. Point-serial correlations between EMQ-R and BRIEF-A subscales and self-reported memory- and concentration problems were investigated (supplementary eTable 1).

To examine the effect of non-response bias, we conducted a sensitivity analysis in randomly selected participants invited to the substudy (n=2090). The non-responders were contacted by telephone (*n* = 343; 56 untested, 237 negative, and 50 positive) and were compared with those who had already responded through electronic reminders (*n* = 1692).

Analyses were performed using Stata (Stata Statistical Software, release 16 and 17, Stata Corp., College Station, TX), R (version 4.2.2), and SPSS 27 (IBM).

### Patient and Public Involvement statement

We have involved patients in the Norwegian Corona Cohort since the spring of 2020. Our focus on persisting cognitive symptoms were initiated after reports of such symptoms by a patient representative which led to the inclusion of relevant questions in the following questionnaires. The study has had a two-way communication with our participants through newsletters and free-text-fields in the questionnaires. We have regular meetings with patients and the patient organization Norwegian Covid Association.

## Results

### Participants and follow-up

Of 120 605 participants, 15 737 were SARS-CoV-2-positive (58% from MSIS and 42% self-report), 67 305 negative, and 37 563 untested at the end of follow-up (July 6, 2022; Supplementary eFigure 1). The follow-up varied between the groups and 710 SARS-CoV-2 positive, 35 309 negative, and 18 997 untested participants had >12 months follow-up after the test (Table 1). In total, 530 200 questionnaires were included in the analyses. Of the participants, 69% were women, 41% had a per household income/ year >1 000 000 NOK (about US$ 100 000), and the mean age was 49 years (SD = 13.7 years). SARS-CoV-2-positive were younger, had a higher income, a lower BMI, and fewer underlying medical conditions, and were less likely to be current smokers compared with negative and untested participants.

**Table 1.** Frequency distribution and mean values of covariates by SARS-CoV-2 status at fourth follow-up^a^ (*n* = 120 605).

|  | **SARS-CoV-2 positive<sup>a</sup>**<br>(*n* = 15 737) | **SARS-CoV-2-negative**<br>(*n* = 67 305) | **Untested<sup>b</sup>**<br>(*n* = 37 563) |
| --- | --- | --- | --- |
| **Characteristics** |  |  |  |
|  | **Median (IQR)<sup>c</sup>** |  |  |
| **Time since test (days)<sup>d</sup>** | 55 (15-106) | 419 (292-501) | 637 (633-669) |
|  | **Mean (SD)** |  |  |
| **Age (years)** | 46 (12.1) | 48 (13.3) | 51 (14.7) |
|  | **n (%)** |  |  |
| 18-29 | 1298 (8) | 5081 (8) | 3150 (8) |
| 30-39 | 3956 (25) | 14 331 (21) | 6490 (17) |
| 40-49 | 4899 (31) | 17 108 (26) | 7769 (21) |
| 50-59 | 3429 (22) | 16 164 (24) | 8765 (24) |
| 60-69 | 1608 (10) | 10 325 (15) | 7166 (19) |
| 70+ | 547 (4) | 4296 (6) | 4223 (11) |
| **Sex** |  |  |  |
| Men | 4287 (27) | 18 879 (28) | 13 750 (37) |
| Women | 11 450 (73) | 48 426 (72) | 23 813 (63) |
| **Income (NOK per household and year)** |  |  |  |
| < 299 999 | 477 (3) | 2422 (4) | 1492 (4) |
| 300 000-599 999 | 2181(14) | 12 185 (18) | 7049 (19) |
| 600 000-1000 000 | 4148 (26) | 19 323 (29) | 9905 (26) |
| >1000 000 | 7096 (45) | 27 681 (41) | 11 890 (32) |
| Missing | 1835 (12) | 5692 (8) | 7227 (19) |
| **Body mass index (kg/ m<sup>2</sup>)** |  |  |  |
| <25 | 7603 (48) | 30 902 (46) | 16 924 (45) |
| ≥25 | 8018 (51) | 35 946 (53) | 20 301 (54) |
| Missing | 116 (1) | 457 (1) | 338 (1) |
| **Smoking status** |  |  |  |
| Never | 8443 (54) | 35 418 (53) | 18 947 (50) |
| Former | 6109 (39) | 25 651 (38) | 14 344 (38) |
| Current | 890 (5) | 4913 (7) | 3313 (9) |
| Missing | 295 (2) | 1323 (2) | 959 (3) |
| **Underlying medical conditions<sup>e</sup>** |  |  |  |
| No | 11 817 (75) | 47 947 (71) | 25 654 (68) |
| Yes | 3777 (24) | 18 622 (28) | 11 393 (31) |
| Missing | 143 (1) | 736 (1) | 516 (1) |

<sup>a</sup> The SARS-CoV-2 status of the negative participants was obtained from the Norwegian Messaging System for Reporting of Infectious Diseases (MSIS) registry. The SARS-CoV-2-positive status was obtained from the MSIS registry 58% or self-report 42%. Only one SARS-CoV-2-positive-test was included for each participant.
<sup>b</sup> Participants that remained untested for the whole follow-up period
<sup>c</sup> Interquartile range.
<sup>d</sup> Time from the first SARS-CoV-2 positive test or first SARS-CoV-2 negative test to last follow up. For participants remaining untested, time from baseline to the last follow-up is shown.
<sup>e</sup> Chronic heart disease, high blood pressure, chronic lung disease (not asthma), asthma, diabetes, receiving immunodeficiency treatment, cancer (under treatment).

### Symptoms among SARS-CoV-2-positive participants

The temporal trajectory of each symptom differed by SARS-CoV-2 status (Unadjusted: Figure 1 and Supplementary eFigure 3, and adjusted: Table 2, Supplementary eTables 2 and 3). When comparing SARS-CoV-2-positive participants with negative participants (1-3 months after a negative test), there were several persistent symptoms. The strongest findings in the adjusted analyses were memory problems (3-6 months: adjusted odds ratio (OR) = 9.1, 95% confidence interval (CI) = 7.5-10.9; 6-12 months: OR = 13.3, CI = 10.7-16.6; 12-18 months: OR = 7.8, CI = 5.7-10.8; >18 months: OR = 9.96, CI = 4.3-23.0), concentration problems (3-6 months: OR = 5.7, CI = 4.9-6.7; 6-12 months: OR = 7.5, CI = 6.1-9.1; 12-18 months: OR = 5.3, CI = 3.9-7.1; >18 months: OR = 4.9, CI = 2.1-11.1), and anosmia and dysgeusia (3-6 months: OR = 5.2, 95% CI = 4.4-6.2, 6-12 months: OR = 8.9, CI = 7.2-11;12-18 months; OR = 5.5, CI = 4.0-7.5; >18 months: OR = 5.3, CI = 2.4-2.1) (Table 2). Self-assessed worsening of overall health was also significantly higher among the SARS-CoV-2-positive participants, with the highest risk being at 18 months (OR = 3.7, CI = 2.0-6.8). The risk of reporting dyspnoea and fatigue was higher for the positive than negative participants, but the risk attenuated over time. SARS-CoV-2 positive participants were equally or less likely to report fever, headache, cough, muscular pain, nasal symptoms, sore throat, and abdominal pain late in follow-up compared to the negative participants (Supplementary eTable 2).

**Figure 1.**
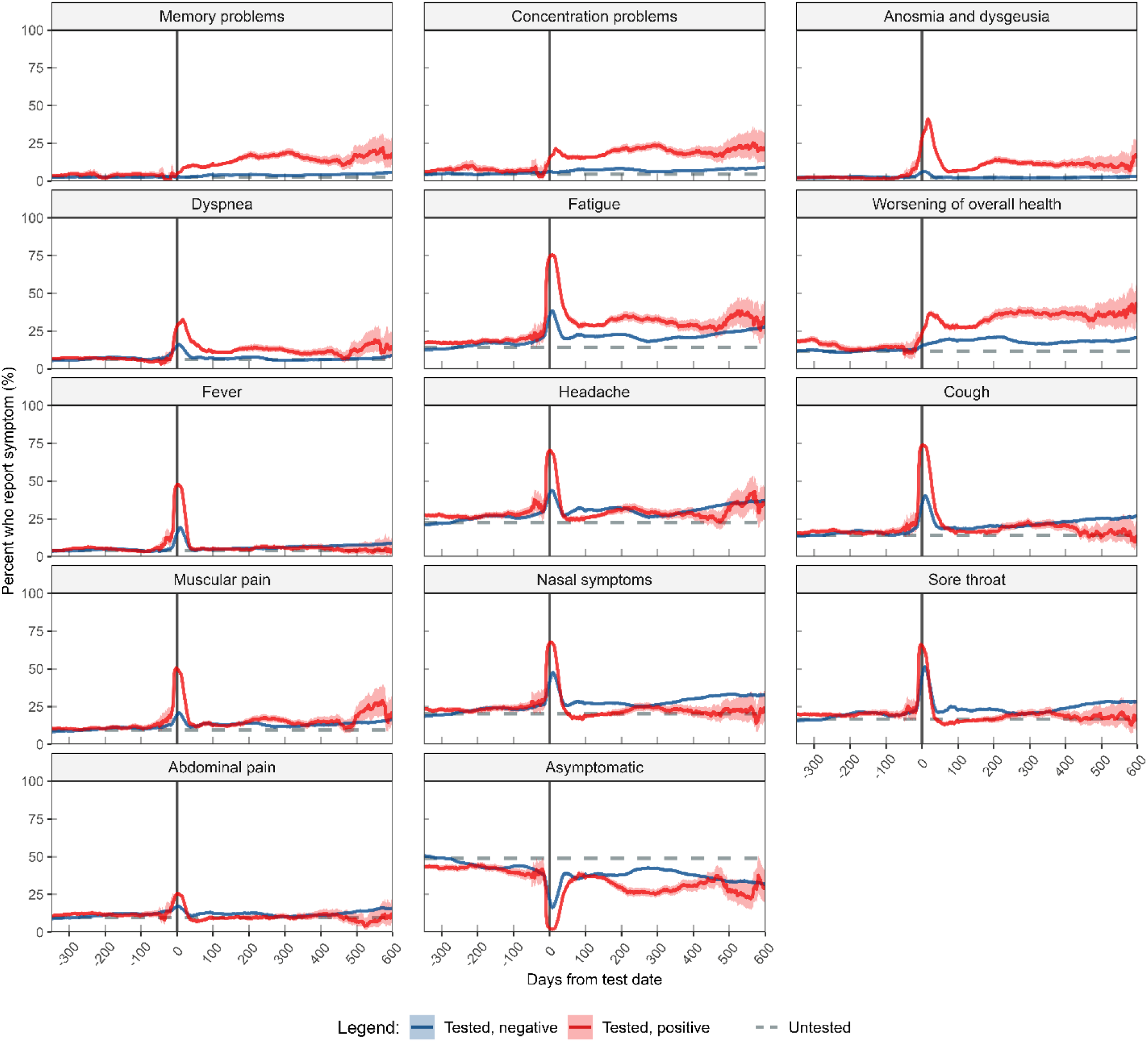
The frequencies (moving average and 95% confidence intervals) of symptoms reported before, during and after a positive or negative SARS-CoV-2 test. Participants are grouped by their SARS-CoV-2 status at the end of follow-up. Participants remaining untested for the complete study period (without a test date) are represented with a horizontal (flat) line representing their average (supplementary eMethods 5). The horizontal grey dashed line represents the mean response for untested study participants. The vertical line represents the day of the positive- or negative SARS-CoV-2 test. The figure is based on all completed questionnaires (N=530 200). In each questionnaire, the presence of a symptom the past three weeks was reported by the participant. Note that the 95% confidence interval for the Tested, negative group falls inside the line and it is therefore not visible.

**Table 2.** Odds ratio (OR) estimates and 95% confidence interval (CI) for reporting each symptom at different time-intervals after a positive or negative SARS-CoV-2 test (or inclusion for untested) with symptoms reported 1-3 months after a negative test as reference (n = 120 605)*.

|  |  | Time since test (months)<sup>#</sup> |  |  |  |  |  |  |  |  |  |  |  |
| --- | --- | --- | --- | --- | --- | --- | --- | --- | --- | --- | --- | --- | --- |
|  |  | 0-1 |  | >1-3 |  | >3-6 |  | >6-12 |  | >12-18 |  | >18 |  |
| Symptoms | SARS-CoV-2 status | Yes: n (%) | OR (95% CI) | Yes: n (%) | OR (95% CI) | Yes: n (%) | OR (95% CI) | Yes: n (%) | OR (95% CI) | Yes: n (%) | OR (95% CI) | Yes: n (%) | OR (95% CI) |
| Memory problems<sup>a</sup> | Untested | -- | 0.8<br>(0.22-2.97) | 4<br>(3.6) | 1.01<br>(0.28-3.65) | 22<br>(2.2) | 0.61<br>(0.36-1.04) | 1502<br>(2.5) | 0.72<br>(0.64-0.81) | 58<br>(4.3) | 2.6<br>(1.81-3.73) | 441<br>(2.4) | 1<br>(0.84-1.19) |
|  | Negative | 253<br>(2.7) | 0.8<br>(0.66-0.96) | 604<br>(3.4) | 1<br>(Ref.) | 720<br>(4.1) | 1.33<br>(1.16-1.54) | 836<br>(4.1) | 1.54<br>(1.34-1.78) | 1289<br>(4.6) | 1.49<br>(1.32-1.69) | 417<br>(6) | 1.98<br>(1.68-2.34) |
|  | Positive | 395<br>(6.8) | 3.34<br>(2.78-4.02) | 502<br>(9.8) | 6.18<br>(5.16-7.42) | 481<br>(12) | 9.05<br>(7.49-10.94) | 339<br>(17.3) | 13.3<br>(10.69-16.55) | 101<br>(13.7) | 7.83<br>(5.68-10.79) | 13<br>(15.5) | 9.96<br>(4.31-23.01) |
| Concentration problems<sup>a</sup> | Untested | -- | 1.46<br>(0.08-25.56) | 7<br>(6) | 0.85<br>(0.29-2.54) | 46<br>(4.5) | 0.71<br>(0.47-1.06) | 2812<br>(4.7) | 0.64<br>(0.58-0.71) | 92<br>(6.8) | 2.34<br>(1.72-3.19) | 698<br>(3.7) | 0.8<br>(0.69-0.92) |
|  | Negative | 586<br>(6) | 0.95<br>(0.82-1.09) | 1276<br>(7) | 1<br>(Ref.) | 1336<br>(7.6) | 1.17<br>(1.04-1.3) | 1490<br>(7.2) | 1.31<br>(1.17-1.47) | 2179<br>(7.5) | 1.16<br>(1.05-1.27) | 672<br>(9.6) | 1.46<br>(1.27-1.68) |
|  | Positive | 1040<br>(17.8) | 6.05<br>(5.23-7) | 830<br>(16.2) | 5.61<br>(4.81-6.54) | 662<br>(16.6) | 5.71<br>(4.85-6.73) | 423<br>(21.6) | 7.46<br>(6.11-9.1) | 141<br>(19) | 5.27<br>(3.93-7.06) | 17<br>(20.2) | 4.86<br>(2.13-11.07) |
| Anosmia and dysgeusia<sup>b</sup> | Untested | 6898<br>(3.5) | 1.01<br>(0.91-1.13) | 495<br>(1.4) | 0.65<br>(0.56-0.74) | 1018<br>(1.3) | 0.58<br>(0.51-0.65) | 564<br>(1) | 0.37<br>(0.33-0.43) | 15<br>(1.1) | 0.73<br>(0.42-1.28) | 258<br>(1.4) | 0.69<br>(0.58-0.81) |
|  | Negative | 929<br>(5.8) | 3.27<br>(2.88-3.71) | 575<br>(2.4) | 1<br>(Ref.) | 398<br>(2) | 0.83<br>(0.72-0.96) | 459<br>(2.2) | 0.96<br>(0.83-1.11) | 704<br>(2.4) | 1.13<br>(0.99-1.28) | 234<br>(3.4) | 1.31<br>(1.09-1.57) |
|  | Positive | 2004<br>(33.8) | 66.22<br>(57.75-75.93) | 587<br>(11.4) | 9.65<br>(8.31-11.21) | 310<br>(7.7) | 5.22<br>(4.38-6.22) | 248<br>(12.6) | 8.88<br>(7.24-10.88) | 80<br>(10.8) | 5.47<br>(4-7.48) | 13<br>(15.5) | 5.28<br>(2.44-11.43) |

Table 2 cont.
|  |  | Time since test (months)<sup>#</sup> |  |  |  |  |  |  |  |  |  |  |  |
| --- | --- | --- | --- | --- | --- | --- | --- | --- | --- | --- | --- | --- | --- |
|  |  | 0-1 |  | >1-3 |  | >3-6 |  | >6-12 |  | >12-18 |  | >18 |  |
| Symptoms | SARS-CoV-2 status | Yes: n (%) | OR (95% CI) | Yes: n (%) | OR (95% CI) | Yes: n (%) | OR (95% CI) | Yes: n (%) | OR (95% CI) | Yes: n (%) | OR (95% CI) | Yes: n (%) | OR (95% CI) |
| Dyspnoea<sup>b</sup> | Untested | 17386 (8.7) | 0.98 (0.91-1.05) | 1445 (4.2) | 0.58 (0.53-0.64) | 2672 (3.5) | 0.48 (0.44-0.51) | 2152 (3.3) | 0.48 (0.44-0.52) | 91 (6.8) | 1.62 (1.22-2.14) | 664 (3.5) | 0.58 (0.52-0.65) |
|  | Negative | 2212 (13.8) | 3.04 (2.78-3.32) | 1761 (7.2) | 1 (Ref.) | 1348 (6.9) | 0.99 (0.9-1.08) | 1380 (6.7) | 0.95 (0.87-1.05) | 1858 (6.4) | 0.99 (0.91-1.07) | 625 (9) | 0.97 (0.86-1.09) |
|  | Positive | 1765 (29.7) | 14.14 (12.69-15.76) | 769 (14.9) | 4.07 (3.6-4.6) | 449 (11.1) | 2.44 (2.12-2.81) | 247 (12.6) | 2.66 (2.2-3.21) | 82 (11.1) | 2.02 (1.49-2.73) | 10 (11.9) | 2.03 (0.9-4.59) |
| Fatigue<sup>b</sup> | Untested | 34532 (17.4) | 0.51 (0.49-0.54) | 3779 (11) | 0.47 (0.44-0.49) | 8332 (10.9) | 0.43 (0.41-0.45) | 7667 (11.7) | 0.57 (0.54-0.6) | 227 (16.9) | 1.43 (1.19-1.73) | 1982 (10.5) | 0.63 (0.59-0.68) |
|  | Negative | 5617 (35) | 2.92 (2.76-3.1) | 5245 (21.5) | 1 (Ref.) | 4306 (22) | 1.08 (1.02-1.15) | 4359 (21) | 1.14 (1.08-1.21) | 6578 (22.8) | 1.23 (1.17-1.3) | 1953 (28) | 1.44 (1.33-1.55) |
|  | Positive | 4236 (71.4) | 27.63 (25.25-30.23) | 1735 (33.5) | 2.86 (2.62-3.12) | 1182 (29.3) | 2.02 (1.83-2.23) | 647 (32.9) | 2.47 (2.16-2.82) | 218 (29.5) | 1.78 (1.44-2.2) | 26 (31) | 1.3 (0.72-2.36) |
| Self-assessed worsening of overall health<sup>c</sup> | Untested | 27 (13.3) | 0.5 (0.3-0.82) | 1625 (11.9) | 0.49 (0.45-0.53) | 8070 (10.6) | 0.42 (0.39-0.44) | 8652 (13.3) | 0.66 (0.62-0.69) | 107 (10.7) | 0.52 (0.4-0.66) | 2137 (11.5) | 0.55 (0.51-0.59) |
|  | Negative | 2308 (16.5) | 0.84 (0.78-0.9) | 4461 (18.7) | 1 (Ref.) | 3740 (19.4) | 1.06 (1-1.13) | 3811 (18.8) | 1.02 (0.96-1.09) | 5193 (18.1) | 1 (0.95-1.06) | 1432 (20.7) | 1.01 (0.93-1.1) |
|  | Positive | 1717 (29.7) | 2.38 (2.18-2.59) | 1537 (30.6) | 2.61 (2.38-2.85) | 1041 (28.0) | 2.32 (2.1-2.57) | 618 (37.1) | 3.63 (3.15-4.19) | 238 (36.2) | 3.44 (2.77-4.26) | 30 (40.0) | 3.77 (2.05-6.94) |

<sup>a</sup> Adjusted for age (10-years categories), gender (men, women) body mass index (<25 kg/ m2, ≥25 kg/ m2), income level per household (< 299 999, 300 000-599 999, 600 000-1000 000, >1000 000 NOK, missing), smoking status (never, former, current, missing), underlying medical condition (no, yes, missing) and symptom status at baseline (no, yes, missing).
<sup>#</sup> For the untested, time since baseline was used in place of time since a positive- or negative SARS-CoV-2 test.
<sup>a</sup> n = 200 778 questionnaires, 120 605 participants
<sup>b</sup> n = 530 200 questionnaires, 120 605 participants
<sup>c</sup> n = 304 958 questionnaires, 120 605 participants

### Symptoms among SARS-CoV-2-negative and untested participants

Eighty percent of participants with a negative test reported at least one symptom indicating an infection in the three-week period around testing (data not shown). At later follow-up periods, SARS-CoV-2-negative participants had a higher risk of reporting symptoms compared to 1-3 months after the negative test. Memory problems (3-6 months: OR = 1.3, CI = 1.2-1.5; >18 months: OR = 2.0, CI = 1.7-2.3), fever (3-6 months: OR = 1.2, CI = 1.00-1.4; >18 months: OR = 2.1, CI = 1.8-2.3), and cough (3-6 months: OR = 1.0, CI = 0.94-1.1; >18 months: OR = 1.5, CI = 1.4-1.7) were increasingly reported over time among negative participants (Table 2 and Supplementary eTable 2). We therefore conducted analyses comparing SARS-CoV-2-positive with negative participants at each time point (Supplementary eTable 3). Untested participants were less likely to report symptoms compared to SARS-CoV-2-negative participants.

### Disease severity and persisting symptoms

A longer time bedridden with COVID-19 was associated with an increased risk of persisting symptoms, except for sore throat and nasal symptoms. The strongest association in the adjusted analyses was seen for memory problems (bedridden 1-6 days: OR = 2.1, CI = 2.0-2.7, bedridden 7-13 days: OR = 5.3, CI = 4.3-6.6, and bedridden >=14 days: OR 9.1, CI 6.5-12.8, compared to not being bedridden during COVID-19) (Supplementary eMethods 6, and eTable 4).

### Sensitivity analyses

When excluding participants with SARS-CoV-2 Omicron variants (BA.1 and BA.2), the results were consistent (eFigure 4), but the ORs for memory problems, concentration problems, and anosmia and dysgeusia 3-6 months after the positive test were higher compared to when all cases were included (data not shown). In other sensitivity analyses, we included only those who completed all questionnaires, excluded hospitalized COVID-19 participants (4%), and participants with self-reported SARS-CoV-2 status, and the results remained largely unchanged (data not shown). In one sensitivity analysis, only participants answering questionnaires both before and after a positive or negative SARS-CoV-2 test were included and the results were similar to when all questionnaires were included (data not shown).

### Validated questionnaires on cognitive function, phone cohort and serology

Compared to SARS-CoV-2 negative participants, positive participants reported more functional memory problems on the EMQ-R (Supplementary eFigure 5; substudy: Mean difference = 0.262, CI = 0.204 - 0.321; fourth follow-up: Mean difference = 0.329, CI = 0.298 - 0.394), and poorer working memory function (BRIEF-A subscale, Supplementary eFigure 6; Mean difference = 0.620, CI= 0.346-0.894). There were no significant differences on the other BRIEF-A subscales. With regard to change in EMQ-R, those who converted to positive at fourth follow-up had larger changes (*n =* 223; M=.124, SD = .63) than those who remained negative or untested (*n* = 2489; M=.029, SD = .449), mean difference = .096, CI = .031, .160, p = .004). We performed correlation tests between EMQ-R/BRIEF-A Working memory subscale, and self-reported memory- and concentration problems and found small (Pearson’s r from 0.26 to 0.40), but significant correlations between these measurements (Supplementary eTable 1).

In the subgroup of participants contacted by telephone (n=343), a lower proportion, reported symptoms compared with those who responded through electronic invitations. This difference was more pronounced among positive than among negative participants (Supplementary eFigure 7).

In the add-on serology study of 966 participants, blood samples were analysed for SARS-CoV-2 antibodies, 3 (0.3%) were positive but classified as negative or untested at the time of sampling (Supplementary eMethods 2).

## Discussion

In this prospective cohort study, we have analysed the temporal trajectory of 14 symptoms (13 symptoms and self-assessed worsening of overall health) before, during and after a positive or negative SARS-CoV-2 test. When comparing SARS-CoV-2-positive participants with negative participants, persistent symptoms were memory- and concentration problems, anosmia and dysgeusia, dyspnoea, fatigue, and self-assessed worsening of overall health. The cognitive symptoms and self-assessed worsening of overall health did not attenuate over time. SARS-CoV-2 positive participants also scored worse than negative participants in validated tests of functional and working memory.

Our finding of memory- and concentration problems as long-COVID symptoms is in line with a report by the WHO^28^ and several studies.^9,10,12–16,29^ However, the results elaborates this association with several important findings. Firstly, the association between COVID-19 and memory- and concentration problems was strong, and after the acute phase – as strong as the association between COVID-19 and anosmia and dysgeusia. Secondly, memory- and concentration problems increased in the 0-6 months interval after a positive SARS-CoV-2 test and persisted throughout the follow-up time up to 22 months. In contrast, the other symptoms peaked in prevalence during the acute phase and thereafter exhibited a downwards trend. To our knowledge, this has not previously been reported and may indicate that the pathophysiological processes underlying the cognitive symptoms may differ from other long-COVID symptoms.

The strong association and a temporal trajectory specific for cognitive symptoms warrants further research.

Several studies have examined the neuropathological effects of the SARS-CoV-2 virus and possible mechanisms associated with COVID-19.^30–33^ Brain imaging studies have reported a greater reduction of grey matter^30^ and an increase of white matter lesions after COVID-19^31^, whereas an autopsy study found the virus persisting in the brain of COVID-19 patients.^32^ Another study has suggested that synaptic signalling of neurons may expand after COVID-19 disease^33^ and could be linked to neurodegenerative and psychiatric disorders.^34–36^ The SARS-CoV-2-positive participants’ scores on BRIEF-A and EMQ-R in the current study indicate that most participants reporting memory- and concentration problems had mild cognitive impairments. However, even slight cognitive impairments can have profound effects on everyday functioning because working memory supports decision-making, navigation, and problem-solving, and it helps keep track of conversations.^37^ The fact that cognitive symptoms are persistent, and most of the population have been, or will be, infected by the SARS-CoV-2 virus, makes it crucial that further research focuses on the pathophysiology, and treatment (including whether immediate treatment upon symptom debut are effective) of these symptoms.^38^

Our findings are consistent with a large recent cohort study examining cognitive trajectories in older adults^29^. They asked about history of COVID-19 in the year before each questionnaire, and a decline in cognition was observed after COVID-19 compared to pre-pandemic year.

A recently published report from the Lifelines Study analysed 23 somatic (not cognitive) self-reported symptoms from 76 000 participants, 90-150 days after COVID-19, and had a matched non-infected control group.^2^ Our findings on anosmia and dysgeusia, dyspnoea, and fatigue are consistent with their findings. However, we included a control group with negative tests (80% of the participants with a negative test reported ≥1 symptoms of an infection at the time of testing), allowing us to identify symptoms specific to the SARS-CoV-2 virus compared to other infections. This could be one of the reasons why the current study did not find any association between COVID-19 and persisting fever, headache, cough, nasal symptoms, sore throat, or abdominal pain when we compared SARS-CoV-2 positive and negative participants. We observed an upwards trend in symptoms associated with upper respiratory tract infections (URTI) among the SARS-CoV-2-negative participants over time. This was probably because lockdown measures were gradually relaxed throughout the pandemic.

Our findings are also consistent with findings from a large cohort study following 138 818 SARS-CoV-2-positive and 5 985 227 controls from the US Department of Veterans Affairs.^39^ In that study, diagnoses were tracked up to two years after COVID-19 and the risk of 27 of 79 diagnoses were still elevated 2 years after COVID-19 compared to non-infected controls. Furthermore, in a cross-sectional study (EPILOC) on 50 457 SARS-CoV-2 positive participants reported that fatigue, neurocognitive impairment, dyspnoea, anosmia, and dysgeusia persisted 6-12 months after the SARS-CoV-2 infection.^40^

The current study is a large prospective cohort with a long follow-up period, and multiple follow-up questionnaires, including before disease, with a high response rate. The study included a SARS-CoV-2-negative control group, making it possible to capture differences between COVID-19 and other infections. The SARS-CoV-2 status was obtained from accredited laboratories through MSIS which covers nearly 100% of the population. Another strength is the proportion of missing data which was < 3% for most of the covariates.

Participants were asked to report symptoms experienced in the last three weeks in each questionnaire, and we cannot rule out recall bias. Knowledge of exposure status and media reports on long-COVID symptoms may have biased the self-reported outcome assessment leading to exaggerated ORs for known long-COVID symptoms, at least later in the follow-up period once these became common knowledge (“nocebo effect”). Since data on the symptoms were reported as a three-week symptom prevalence, participants reporting “no symptoms” could still have had symptoms at some point in time. Our coverage of long-COVID symptoms is not exhaustive, and symptoms not known to be related to acute or long-COVID early in the pandemic are not included (i.e., orthostatic intolerance, dysautonomia, palpitations, light-headedness, or post-exertional malaise). There may be testing bias in the comparison between the tested and untested participants. For this reason, we provided results both on positive, negative and untested participants.

We cannot rule out non-differential measurement error caused by false negative SARS-CoV-2 tests. However, in our add-on study with serological screening, we only identified 3 out of 966 (0.3%) unacknowledged cases of COVID-19. In the sensitivity analysis of the substudy, we contacted a subgroup of participants by telephone, and found that a lower proportion reported symptoms compared with those who responded through electronic invitations. This difference was more pronounced among SARS-CoV-2-positive, than among negative participants and could have led to an overestimation of the effect of COVID-19 on symptoms. We did not adjust for multiple testing in the statistical analyses. Nevertheless, the clinical relevance of the actual differences and sizes is the most essential.

The generalizability of our findings is limited by our cohort composition, which has lower proportions of males, non-white ethnic groups, individuals with lower income, older individuals and no children compared to the Norwegian population. Persistent long-COVID symptoms lasting up to 22 months were memory- and concentration problems, anosmia and dysgeusia, dyspnoea, fatigue and self-assessed worsening of overall health. We could not establish an association between COVID-19 and persisting fever, headache, cough, nasal symptoms, sore throat, or abdominal pain suggesting that the role of previous COVID-19 infections in patients presenting with such symptoms may not be important.

The core long-COVID-19 symptoms may have a long-lasting negative impact on peoples’ lives and should be in focus in further research and in long-COVID rehabilitation.

## Acknowledgments

We thank Dr Fridtjof Lund Johansen at Oslo University Hospital for organizing and carrying out the serological analyses.

## Author contributions

Merete Ellingjord-Dale, Anders B. Nygaard, and Arne Søraas had full access to all the data in the study and take responsibility for the integrity of the data and the accuracy of the data analysis.

Merete Ellingjord-Dale and Anders B. Nygaard contributed equally as co-first authors.

Conceptualization: Arne Søraas and Karl Trygve Kalleberg.

Design of study and analyses: Merete Ellingjord-Dale, Anders B. Nygaard, Karl Trygve Kalleberg, John Arne Dahl, Sonja H. Brunvoll, Nathalie C. Støer, Linda Geng, Giske Ursin and Arne Søraas.

Acquisition, analysis, or interpretation of data: all authors.

Data curation and statistical analysis: Merete Ellingjord-Dale, Anders B. Nygaard, Ragnhild Bø, Nils Inge Landrø and Arne Søraas.

Drafting of the manuscript: Merete Ellingjord-Dale, Anders B. Nygaard, Sonja H. Brunvoll, Nathalie C. Støer, Giske Ursin, and Arne Søraas.

Critical revision of the manuscript for important intellectual content: all authors.

Administrative and technical support: Mette S. Istre.

Supervision: Giske Ursin, Nathalie C. Støer and Arne Søraas.

Final approval of the manuscript and work: all authors.

## Additional contributions

The authors would like to thank all the participants in the study.

## Transparancy statement

The lead authors (MED and ABN) affirm that this manuscript is an honest, accurate, and transparent account of the trial being reported; that no important aspects of the study have been omitted; and that any discrepancies from the study as planned, and registered have been explained.

## Data availability statement

Individual level data from the study for the purposes outlined in the consent form can be shared with other researchers in a timely fashion. The data are regulated under the European GDPR regulative and sharing of data must be approved by the Data Protection Officer at Oslo University Hospital. Data will be made available for researchers whose proposed use of the data has been approved.

