## Supplements for "Temporal trajectories of COVID-19 symptoms in adults with 22 months follow-up in a prospective cohort study in Norway"

This online-only material has been provided by the authors to give readers additional information about the work, and it is original and not previously published.

#### Supplementary eFigure 1. Flow chart of the study population.

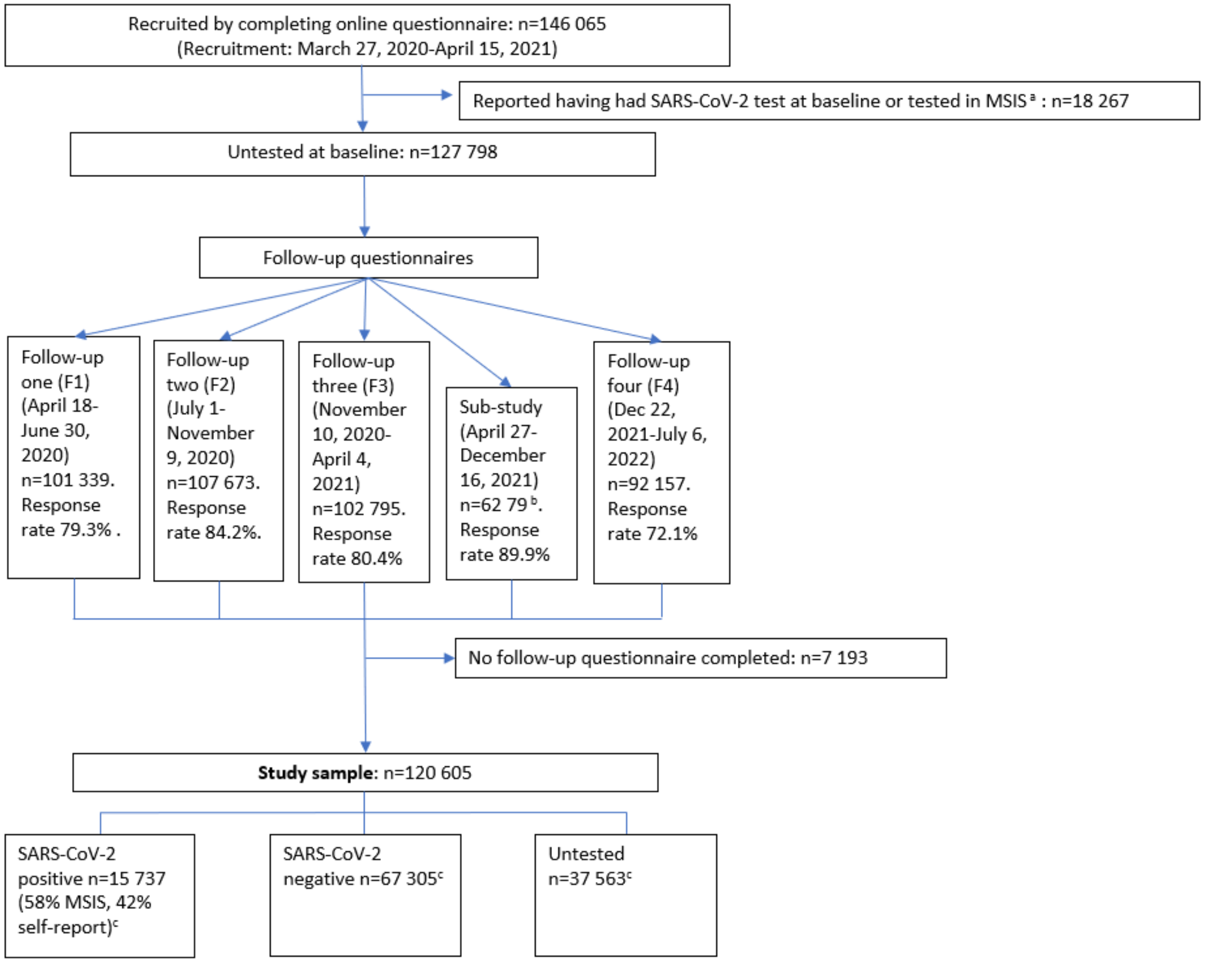

**eFigure 1.** Flow chart of the study population. [a]A sub-study of 6983 participants including all SARS-CoV-2 positive and randomly selected SARS-CoV-2 negative and untested participants. [b]The Norwegian Surveillance System for Communicable Diseases registry (MSIS). [c]SARS-CoV-2 status was obtained from The Norwegian Surveillance System for Communicable Diseases registry or self-report for the SARS-CoV-2 positive participants. SARS-CoV-2 negative status was obtained from the registry only. The numbers of SARS-CoV-2 positive, negative and untested are at the end of follow-up.

#### Supplementary eFigure 2. Proportion of new positive SARS-CoV-2 tests in Norway over time and as test criteria changed.

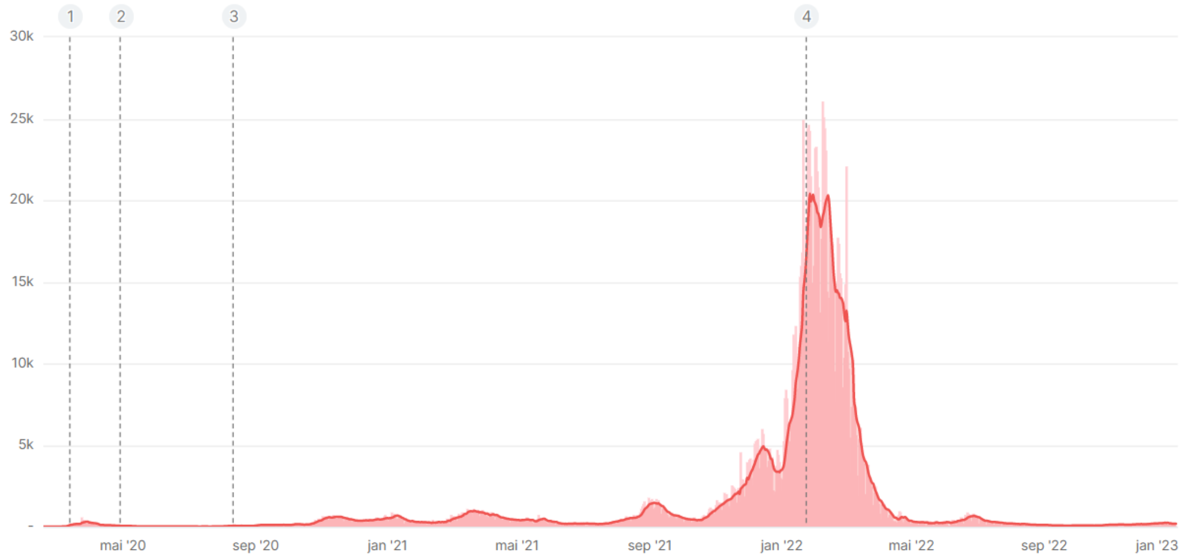

**eFigure 2.** Proportion of new positive SARS-CoV-2 tests in Norway over time and as test criteria changed. 1: March 13, 2020: Testing was reserved for certain patient groups and health care workers with respiratory symptoms. 2: April 29, 2020: Testing was extended to include everyone that a doctor suspected to have been infected by SARS-CoV-2. 3: August 12, 2020: Testing was even further extended and medical evaluation is no longer a requirement to be tested. 4: January 24, 2022. People who have received a 3rd dose of vaccine or have two doses and have had infection in the last three months were no longer offered a PCR test after a positive self-test. (Source: The Norwegian Institute of Public Health).

#### Supplementary eFigure 3. Differences in proportion of participants with symptoms between SARS-CoV-2 positive and negative participants.

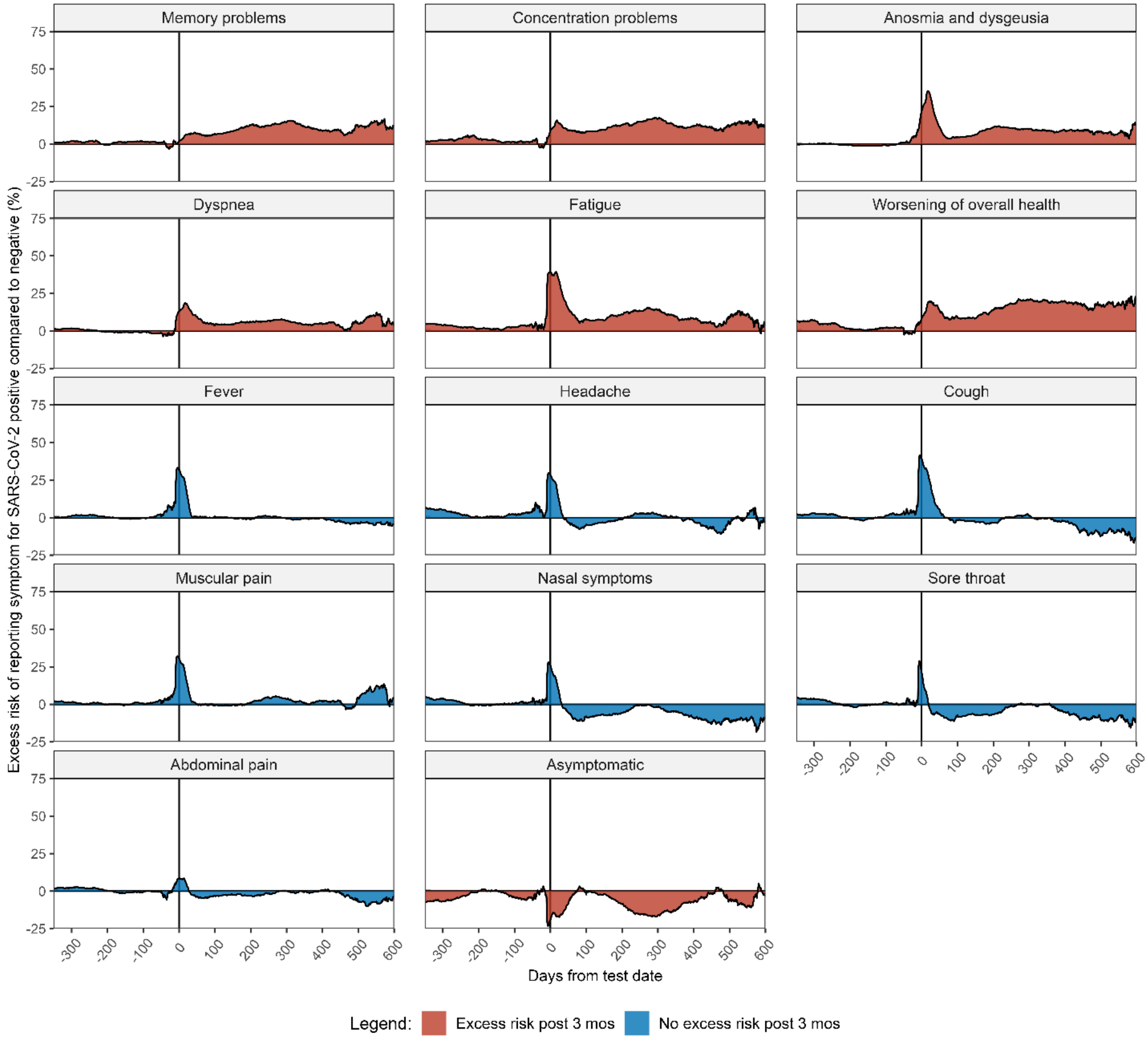

**eFigure 3.** Differences in proportion of participants with symptoms between SARS-CoV-2 positive and negative participants[a] (moving mean difference between the two groups by days from test date[b], n=120 605). Method is described in supplementary eMethods 5.

[a]In the interval -50 to 100 days, the lines represent a 21-day moving average and between <-50 and >100, the lines represent an 84-day moving average.
[b]Time from a SARS-CoV-2 test to a symptom reported (questionnaire date).

#### Supplementary eFigure 4. Symptom trajectories without omicron.

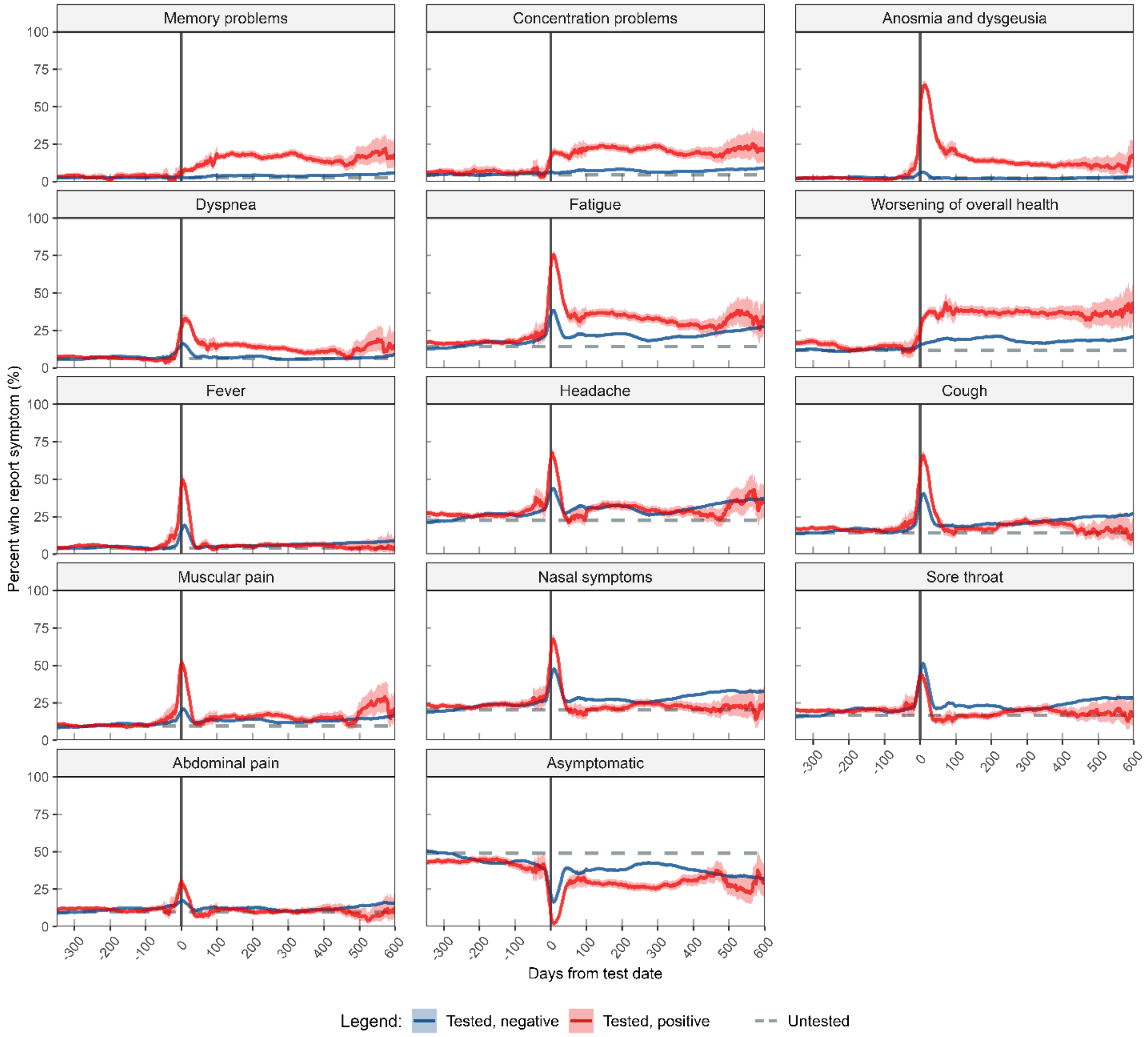

**eFigure 4.** The frequencies (moving average and 95% confidence intervals) of symptoms before, during and after a positive or negative SARS-CoV-2 test before omicron. Participants are grouped by their SARS-CoV-2 status at the end of follow-up, except participants testing positive during the omicron wave (after December 12th 2021) who are omitted. Untested participants (without a test date) are represented with a horizontal (flat) line representing their average. Method is described in supplementary eMethods 5.

#### Supplementary eFigure 5. Everyday Memory Questionnaire by SARS-CoV-2 status.

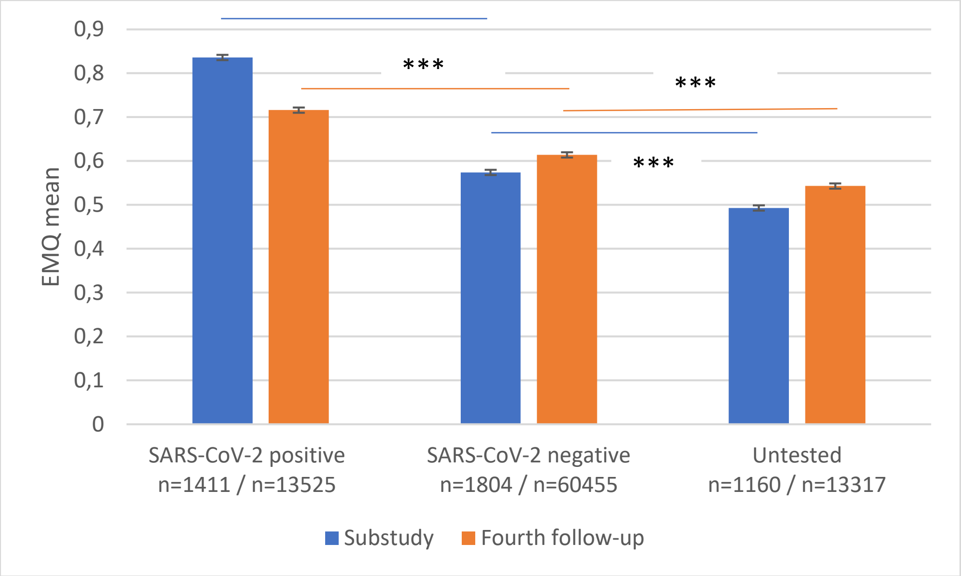

**eFigure 5.** Everyday Memory Questionnaire by SARS-CoV-2 status in the substudy and fourth follow-up. Higher scores represent greater impairments. The bar chart shows the estimated marginal means and error bars +/ -1 SE. *** p<0.001.

**Supplementary eFigure 6. Behavior Rating Inventory of Executive Functions Adult Version (BRIEF-A) Working memory Scores as a Function of SARS-CoV-2 Status.**

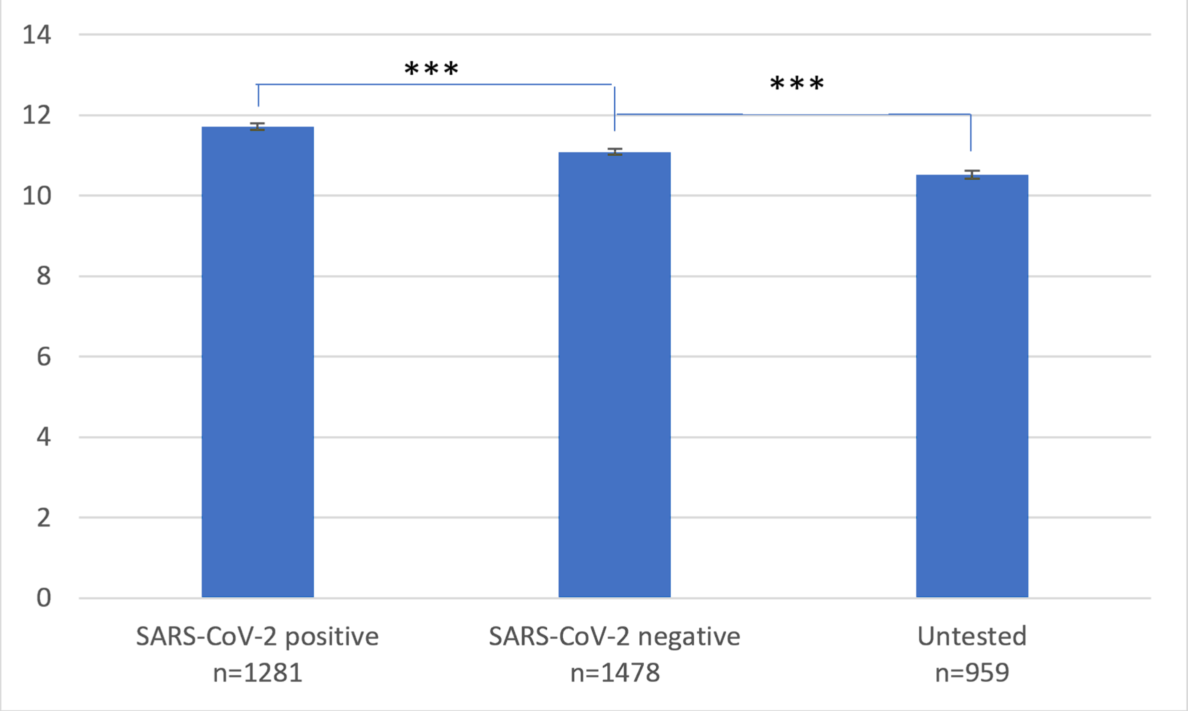

**eFigure 6.** Behavior Rating Inventory of Executive Functions Adult Version (BRIEF-A) Working memory scores as a function of SARS-CoV-2 status in the substudy. Higher scores represent greater impairments. The bar chart shows the estimated marginal means and error bars +/ -1 SD. ***p<0.001.

#### Supplementary eFigure 7. Proportion of participants reporting symptoms by response time in the substudy[a].

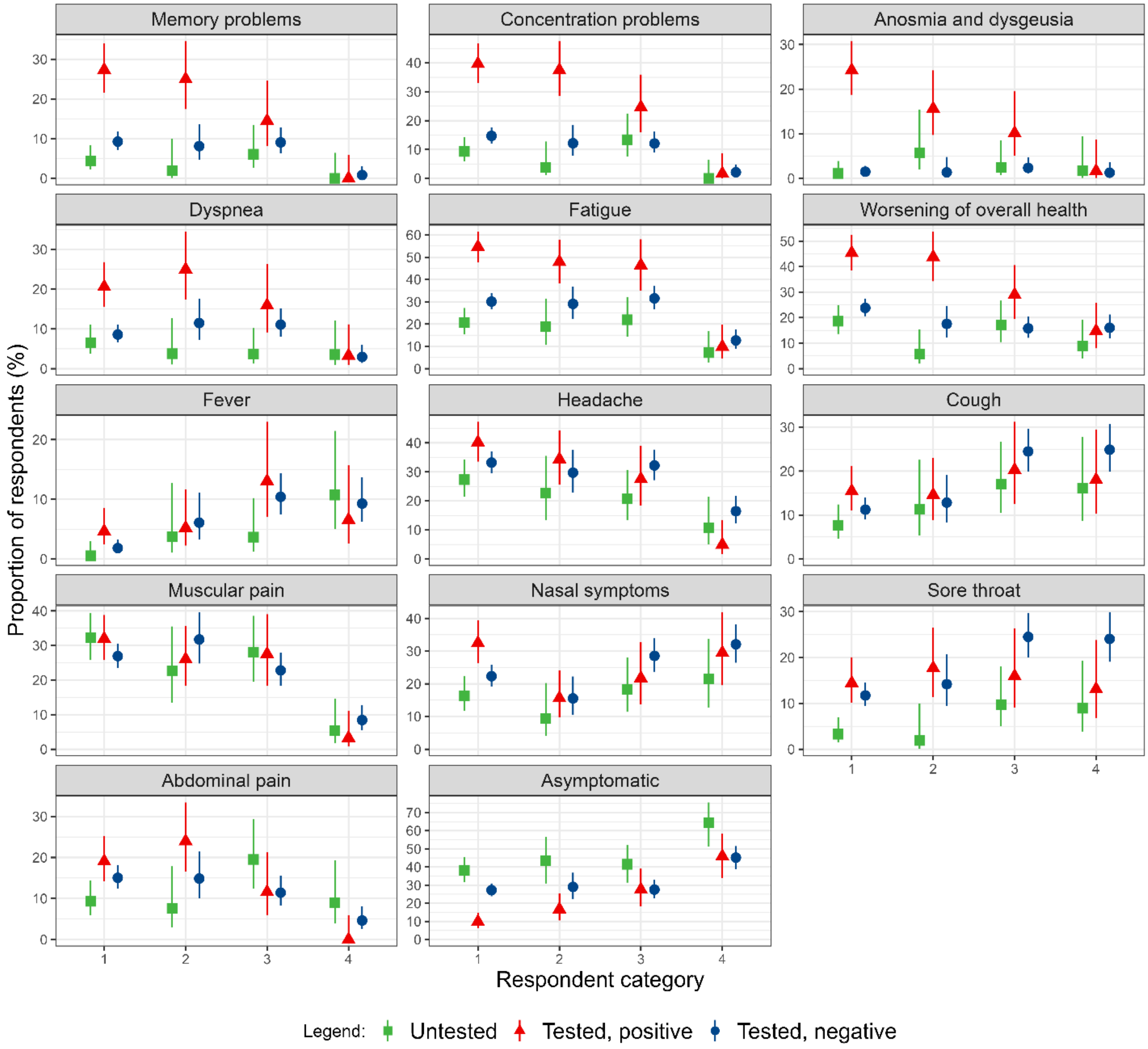

**eFigure 7.** Proportion of respondents reporting having had each symptom during the past three weeks before responding to the substudy questionnaire. The respondents are grouped by the date of response in relation to the initial release of the substudy questionnaire (April 27th, 2021): 1 = early response (Untested n: 183, Positive n: 173, Negative n: 604), 2 = late response (Untested n: 53, Positive n: 90, Negative n: 148), 3 = late response to a shortened questionnaire (Untested n: 82, Positive n: 61, Negative n: 298), 4 = very late response after phone call (Untested n: 56, Positive n: 50, Negative n: 237). Not shown are non-respondents (Untested n: 33, Positive n: 16, Negative n: 36).

#### Supplementary eTable 1. Correlation Between Cognitive Impairments and Self-Reported Memory- And Concentration Problems.

| | **WM**[a] | **EMQ**[b] |
|---|---|---|
| F3 Concentration | .321 *** | .402*** |
| F3 Memory | .256*** | .359*** |
| F4 Concentration | | .357*** |
| F4 Memory | | .378*** |

[a] WM = Working Memory (substudy).
[b] EMQ = Everyday Memory Questionnaire (substudy and fourth follow-up).
*** *p*<.001

**Supplementary eTable 2. Odds ratios (ORs) and 95% confidence intervals (CI) of various symptoms in SARS-CoV-2-untested, negative and positive by time since test as compared with participants who tested negative 1-3 months after a negative test.**

| **Fever (n=530 200, 120 605 participants)** | | | | | | | | | | | | | | | | | | |
|---|---|---|---|---|---|---|---|---|---|---|---|---|---|---|---|---|---|---|
| Time since test (months)# | | | | | | | | | | | | | | | | | | |
| **SARS-CoV-2 status** | 0-1 | | | >1-3 | | | >3-6 | | | >6-12 | | | >12-18 | | | >18 | | |
| | Yes n/ % | OR[c] | 95% CI | Yes n/ % | OR | 95% CI | Yes n/ % | OR | 95% CI | Yes n/ % | OR | 95% CI | Yes n/ % | OR | 95% CI | Yes n/ % | OR | 95% CI |
| Untested | 12595/ 6.3 | 0.59 | 0.54-0.63 | 488/ 1.40 | 0.31 | 0.28-0.35 | 1133/ 1.5 | 0.31 | 0.28-0.34 | 890/ 1.4 | 0.31 | 0.28-0.34 | 33/ 2.5 | 0.72 | 0.5-1.05 | 611/ 3.3 | 0.91 | 0.82-1.01 |
| Negative | 2655/ 16.5 | 5.28 | 4.86-5.73 | 1152/ 4.7 | **1** | **Ref** | 995/ 5.1 | 1.10 | 1.00-1.21 | 1247/ 6.0 | 1.44 | 1.32-1.58 | 2099/ 7.3 | 1.79 | 1.65-1.94 | 628/ 9.0 | 2.05 | 1.83-2.30 |
| Positive | 2403/ 40.5 | 24.79 | 22.46-27.36 | 261/ 5.0 | 1.17 | 1.01-1.37 | 210/ 5.2 | 1.17 | 0.99-1.38 | 116/ 5.9 | 1.35 | 1.09-1.69 | 39/ 5.3 | 1.09 | 0.76-1.56 | 5/ 6.0 | 1.10 | 0.41-2.97 |
| **Headache (n=530 200, 120 605 participants)** | | | | | | | | | | | | | | | | | | |
| Time since test (months)# | | | | | | | | | | | | | | | | | | |
| **SARS-CoV-2 status** | 0-1 | | | >1-3 | | | >3-6 | | | >6-12 | | | >12-18 | | | >18 | | |
| | Yes n/ % | OR | 95% CI | Yes n/ % | OR | 95% CI | Yes n/ % | OR | 95% CI | Yes n/ % | OR | 95% CI | Yes n/ % | OR | 95% CI | Yes n/ % | OR | 95% CI |
| Untested | 52632/ 26.5 | 0.62 | 0.59-0.64 | 5550/ 16.2 | 0.47 | 0.45-0.50 | 13563/ 17.7 | 0.48 | 0.46-0.5 | 14100/ 21.5 | 0.84 | 0.80-0.88 | 274 / 20.4 | 1.22 | 1.02-1.45 | 2887/ 15.4 | 0.64 | 0.61-0.69 |
| Negative | 6566/ 40.9 | 2.15 | 2.04-2.28 | 7364/ 30.2 | **1** | **Ref** | 6173/ 31.5 | 1.16 | 1.1-1.23 | 6026/ 29.1 | 1.15 | 1.09-1.21 | 9560/ 33.1 | 1.28 | 1.22-1.34 | 2578/ 36.9 | 1.49 | 1.38-1.60 |
| Positive | 3703/ 62.4 | 7.18 | 6.63-7.77 | 1311/ 25.3 | 0.80 | 0.73-0.88 | 1100/ 27.2 | 0.85 | 0.77-0.94 | 590/ 30.0 | 1.04 | 0.91-1.18 | 207/ 28.1 | 0.94 | 0.76-1.15 | 32/ 38.1 | 1.39 | 0.8-2.44 |

**eTable 2 cont.**

| **Cough (n=530 200, 120 605 participants)** | | | | | | | | | | | | | | | | | | |
|---|---|---|---|---|---|---|---|---|---|---|---|---|---|---|---|---|---|---|
| Time since test (months)# | | | | | | | | | | | | | | | | | | |
| **SARS-CoV-2 status** | 0-1 | | | >1-3 | | | >3-6 | | | >6-12 | | | >12-18 | | | >18 | | |
| | Yes n/ % | OR | 95% CI | Yes n/ % | OR | 95% CI | Yes n/ % | OR | 95% CI | Yes n/ % | OR | 95% CI | Yes n/ % | OR | 95% CI | Yes n/ % | OR | 95% CI |
| Untested | 37796/ 19 | 0.61 | 0.58-0.64 | 3007/ 8.8 | 0.38 | 0.36-0.41 | 6841/ 8.9 | 0.37 | 0.36-0.39 | 5817/ 8.9 | 0.41 | 0.39-0.43 | 127/ 9.4 | 0.56 | 0.45-0.70 | 2506/ 13.3 | 0.82 | 0.77-0.87 |
| Negative | 5985/ 37.2 | 3.73 | 3.52-3.96 | 4705/ 19.3 | **1** | **Ref** | 3689/ 18.8 | 1.00 | 0.94-1.06 | 4305/ 20.8 | 1.26 | 1.19-1.33 | 7011/ 24.2 | 1.61 | 1.53-1.69 | 1866/ 26.7 | 1.54 | 1.43-1.66 |
| Positive | 4040/ 68.1 | 22.57 | 20.76-24.54 | 1136/ 22.0 | 1.45 | 1.32-1.59 | 661/ 16.4 | 0.88 | 0.79-0.98 | 397/ 20.2 | 1.23 | 1.07-1.42 | 140/ 19 | 1.12 | 0.89-1.41 | 10/ 11.9 | 0.61 | 0.28-1.31 |

| **Muscular pain (n=530 200, 120 605 participants)** | | | | | | | | | | | | | | | | | | |
|---|---|---|---|---|---|---|---|---|---|---|---|---|---|---|---|---|---|---|
| Time since test (months)# | | | | | | | | | | | | | | | | | | |
| **SARS-CoV-2 status** | 0-1 | | | >1-3 | | | >3-6 | | | >6-12 | | | >12-18 | | | >18 | | |
| | Yes n% | OR | 95% CI | Yes n/ % | OR | 95% CI | Yes n/ % | OR | 95% CI | Yes n/ % | OR | 95% CI | Yes n/ % | OR | 95% CI | Yes n/ % | OR | 95% CI |
| Untested | 21711/ 10.9 | 0.61 | 0.57-0.64 | 2643/ 7.70 | 0.56 | 0.52-0.60 | 5587/ 7.3 | 0.53 | 0.50-0.56 | 6059/ 9.3 | 0.82 | 0.78-0.88 | 265/ 19.7 | 3.38 | 2.78-4.1 | 1497/ 8.0 | 0.67 | 0.62-0.73 |
| Negative | 2924/ 18.2 | 2.02 | 1.88-2.18 | 3083/ 12.6 | **1** | **Ref** | 2574/ 13.1 | 1.11 | 1.03-1.20 | 2699/ 13.0 | 1.17 | 1.09-1.26 | 3949/ 13.7 | 1.21 | 1.13-1.29 | 1176/ 16.8 | 1.38 | 1.25-1.52 |
| Positive | 2476/ 41.7 | 13.35 | 12.15-14.68 | 674/ 13.0 | 1.19 | 1.06-1.34 | 499/ 12.4 | 1.22 | 1.07-1.39 | 307/ 15.6 | 1.57 | 1.32-1.86 | 115/ 15.6 | 1.44 | 1.11-1.88 | 17/ 20.2 | 1.50 | 0.76-2.97 |

**eTable 2 cont.**

| **Nasal symptoms (n=530 200, 120 605 participants)** | | | | | | | | | | | | | | | | | | |
|---|---|---|---|---|---|---|---|---|---|---|---|---|---|---|---|---|---|---|
| Time since test (months)# | | | | | | | | | | | | | | | | | | |
| **SARS-CoV-2 status** | 0-1 | | | >1-3 | | | >3-6 | | | >6-12 | | | >12-18 | | | >18 | | |
| | Yes n/ % | OR | 95% CI | Yes n/ % | OR | 95% CI | Yes n/ % | OR | 95% CI | Yes n/ % | OR | 95% CI | Yes n/ % | OR | 95% CI | Yes n/ % | OR | 95% CI |
| Untested | 47174/ 23.7 | 0.64 | 0.61-0.66 | 5372/ 15.7 | 0.52 | 0.49-0.55 | 13401/ 17.5 | 0.56 | 0.54-0.58 | 10788/ 16.5 | 0.60 | 0.57-0.62 | 183/ 13.6 | 0.64 | 0.53-0.77 | 3226/ 17.2 | 0.80 | 0.75-0.84 |
| Negative | 7221/ 44.9 | 3.07 | 2.91-3.24 | 6735/ 27.6 | **1** | **Ref** | 5396/ 27.5 | 1.05 | 1-1.11 | 5512/ 26.6 | 1.13 | 1.08-1.19 | 9194/ 31.8 | 1.39 | 1.32-1.45 | 2248/ 32.2 | 1.31 | 1.22-1.41 |
| Positive | 3632/ 61.2 | 7.29 | 6.76-7.87 | 1021/ 19.7 | 0.63 | 0.58-0.69 | 798/ 19.8 | 0.58 | 0.53-0.65 | 460/ 23.4 | 0.85 | 0.74-0.97 | 160/ 21.7 | 0.70 | 0.57-0.87 | 19/ 22.6 | 0.65 | 0.35-1.21 |

| **Sore throat (n=530 200, 120 605 participants)** | | | | | | | | | | | | | | | | | | |
|---|---|---|---|---|---|---|---|---|---|---|---|---|---|---|---|---|---|---|
| Time since test (months)# | | | | | | | | | | | | | | | | | | |
| **SARS-CoV-2 status** | 0-1 | | | >1-3 | | | >3-6 | | | >6-12 | | | >12-18 | | | >18 | | |
| | Yes n/ % | OR | 95% CI | Yes n/ % | OR | 95% CI | Yes n/ % | OR | 95% CI | Yes n/ % | OR | 95% CI | Yes n/ % | OR | 95% CI | Yes n/ % | OR | 95% CI |
| Untested | 43638/ 21.9 | 0.59 | 0.56-0.61 | 3696/ 10.8 | 0.44 | 0.42-0.47 | 9594/ 12.5 | 0.49 | 0.47-0.51 | 7328/ 11.2 | 0.50 | 0.47-0.52 | 105/ 7.8 | 0.46 | 0.36-0.57 | 2097/ 11.2 | 0.63 | 0.59-0.67 |
| Negative | 7499/ 46.7 | 4.50 | 4.26-4.75 | 5642/ 23.1 | **1** | **Ref** | 4533/ 23.1 | 1.03 | 0.98-1.09 | 4510/ 21.8 | 1.07 | 1.02-1.13 | 7552/ 26.1 | 1.31 | 1.25-1.37 | 1967/ 28.2 | 1.29 | 1.20-1.38 |
| Positive | 3203/ 54.0 | 6.25 | 5.8-6.74 | 761/ 14.7 | 0.56 | 0.51-0.62 | 635/ 15.7 | 0.57 | 0.51-0.63 | 379/ 19.3 | 0.85 | 0.74-0.98 | 140/ 19.0 | 0.84 | 0.67-1.05 | 16/ 19.1 | 0.92 | 0.48-1.76 |

**eTable 2 cont.**

| **Abdominal pain (n=530 200, 120 605 participants)** | | | | | | | | | | | | | | | | | | |
|---|---|---|---|---|---|---|---|---|---|---|---|---|---|---|---|---|---|---|
| Time since test (months)# | | | | | | | | | | | | | | | | | | |
| **SARS-CoV-2 status** | 0-1 | | | >1-3 | | | >3-6 | | | >6-12 | | | >12-18 | | | >18 | | |
| | Yes n/ % | OR[c] | 95% CI | Yes n/ % | OR | 95% CI | Yes n/ % | OR | 95% CI | Yes n/ % | OR | 95% CI | Yes n/ % | OR | 95% CI | Yes n/ % | OR | 95% CI |
| Untested | 23707/ 11.9 | 0.81 | 0.76-0.85 | 2693/ 7.9 | 0.71 | 0.67-0.76 | 6250/ 8.2 | 0.73 | 0.69-0.77 | 5035/ 7.7 | 0.77 | 0.73-0.82 | 127/ 9.4 | 1.30 | 1.04-1.63 | 1171/ 6.2 | 0.69 | 0.64-0.75 |
| Negative | 2479/ 15.4 | 1.57 | 1.46-1.68 | 2944/ 12.1 | **1** | **Ref** | 2402/ 12.3 | 1.06 | 0.99-1.14 | 2405/ 11.6 | 1.1 | 1.02-1.17 | 3622/ 12.5 | 1.14 | 1.07-1.21 | 1082/ 15.5 | 1.29 | 1.17-1.41 |
| Positive | 1319/ 22.2 | 2.86 | 2.61-3.14 | 416/ 8 | 0.63 | 0.56-0.72 | 377/ 9.3 | 0.75 | 0.66-0.86 | 192/ 9.8 | 0.75 | 0.62-0.91 | 80/ 10.8 | 0.83 | 0.62-1.1 | 10/ 11.9 | 0.65 | 0.29-1.43 |
| **Asymptomatic (n=530 200, 120 605 participants)** | | | | | | | | | | | | | | | | | | |
| Time since test (months)# | | | | | | | | | | | | | | | | | | |
| **SARS-CoV-2 status** | 0-1 | | | >1-3 | | | >3-6 | | | >6-12 | | | >12-18 | | | >18 | | |
| | Yes n/ % | OR | 95% CI | Yes n/ % | OR | 95% CI | Yes n/ % | OR | 95% CI | Yes n/ % | OR | 95% CI | Yes n/ % | OR | 95% CI | Yes n/ % | OR | 95% CI |
| Untested | 84733/ 42.6 | 1.72 | 1.66-1.78 | 19835/ 57.8 | 2.33 | 2.23-2.43 | 43031/ 56.2 | 2.36 | 2.27-2.45 | 34403/ 52.5 | 1.55 | 1.49-1.61 | 614/ 45.6 | 0.68 | 0.59-0.79 | 11097/ 59.0 | 1.74 | 1.65-1.83 |
| Negative | 3138/ 19.5 | 0.27 | 0.26-0.29 | 8945/ 36.7 | **1** | **Ref** | 7387/ 37.7 | 1.00 | 0.95-1.05 | 8313/ 40.0 | 0.98 | 0.93-1.03 | 10301/ 35.6 | 0.89 | 0.85-0.93 | 2263/ 32.4 | 0.87 | 0.81-0.94 |
| Positive | 225/ 3.8 | 0.03 | 0.03-0.04 | 1704/ 32.9 | 0.66 | 0.61-0.72 | 1455/ 36 | 0.88 | 0.8-0.96 | 533/ 27.1 | 0.50 | 0.44-0.57 | 227/ 30.8 | 0.69 | 0.56-0.84 | 25/ 29.8 | 0.82 | 0.45-1.47 |

* Adjusted for age (10-years categories), gender (men, women) body mass index (<25 kg/ $m^2$, ≥25 kg/ $m^2$), income level per household (< 299 999, 300 000-599 999, 600 000-1000 000, >1000 000 NOK, missing), smoking status (never, former, current, missing), underlying medical condition (no, yes, missing) and symptom status at baseline (no, yes, missing).

### For the untested, time since baseline was used in place of time since a positive- or negative SARS-CoV-2 test.

#### Supplementary eTable 3. Odds ratios (ORs) and 95% confidence intervals (CI) of various symptoms in SARS-CoV2- untested and SARS-CoV2 positive participants by time since test as compared with participants who tested negative.

**Memory problems (n=200 778, 120 605 participants)**

**Time since test (months)#**

| **SARS-CoV-2 status** | 0-1 | | | >1-3 | | | >3-6 | | | >6-12 | | | >12-18 | | | >18 | | |
|---|---|---|---|---|---|---|---|---|---|---|---|---|---|---|---|---|---|---|
| | Yes n/ % | OR | 95% CI | Yes n/ % | OR | 95% CI | Yes n/ % | OR | 95% CI | Yes n/ % | OR | 95% CI | Yes n/ % | OR | 95% CI | Yes n/ % | OR | 95% CI |
| Untested | - | - | - | 4/ 3.6 | 1.01 | 0.28-3.65 | 22/ 2.2 | 0.46 | 0.27-0.78 | 1502/ 2.5 | 0.47 | 0.42-0.53 | 58/ 4.3 | 1.74 | 1.21-2.5 | 441/ 2.4 | 0.51 | 0.42-0.6 |
| Negative | 253/ 2.7 | 1 | Ref | 604/ 3.4 | 1 | Ref | 720/ 4.1 | 1 | Ref | 836/ 4.1 | 1 | Ref | 1289/ 4.6 | 1 | Ref | 417/ 6.0 | 1 | Ref |
| Positive | 395/ 6.8 | 4.18 | 3.48-5.03 | 502/ 9.8 | 6.18 | 5.16-7.42 | 481/ 12.0 | 6.8 | 5.63-8.23 | 339/ 17.3 | 8.64 | 6.94-10.75 | 101/ 13.7 | 5.26 | 3.81-7.24 | 13/ 15.5 | 5.03 | 2.18-11.62 |

**Concentration problems (n=200 778, 120 605 participants)**

Time since test (months)#

| **SARS-CoV-2 status** | 0-1 | | | >1-3 | | | >3-6 | | | >6-12 | | | >12-18 | | | >18 | | |
|---|---|---|---|---|---|---|---|---|---|---|---|---|---|---|---|---|---|---|
| | Yes n/ % | OR | 95% CI | Yes n/ % | OR | 95% CI | Yes n/ % | OR | 95% CI | Yes n/ % | OR | 95% CI | Yes n/ % | OR | 95% CI | Yes n/ % | OR | 95% CI |
| Untested | - | - | - | 7/ 6.0 | 0.85 | 0.29-2.54 | 46/ 4.5 | 0.61 | 0.4-0.91 | 2812/ 4.7 | 0.49 | 0.44-0.54 | 92/ 6.8 | 2.02 | 1.48-2.75 | 698/ 3.7 | 0.55 | 0.47-0.63 |
| Negative | 586/ 6.0 | 1 | Ref | 1276/ 7.0 | 1 | Ref | 1336/ 7.6 | 1 | Ref | 1490/ 7.2 | 1 | Ref | 2179/ 7.5 | 1 | Ref | 672/ 9.6 | 1 | Ref |
| Positive | 1040/ 17.8 | 6.37 | 5.26-7.37 | 830/ 16.2 | 5.61 | 4.81-6.54 | 662/ 16.6 | 4.88 | 4.15-5.75 | 423/ 21.6 | 5.69 | 4.66-6.94 | 141/ 19.0 | 4.54 | 3.39-6.09 | 17/ 20.2 | 3.33 | 1.46-7.58 |

**eTable 3 cont.**

| **Anosmia and dysgeusia (n=530 200, 120 605 participants)** | | | | | | | | | | | | | | | | | | |
|---|---|---|---|---|---|---|---|---|---|---|---|---|---|---|---|---|---|---|
| Time since test (months)# | | | | | | | | | | | | | | | | | | |
| **SARS-CoV-2 status** | 0-1 | | | >1-3 | | | >3-6 | | | >6-12 | | | >12-18 | | | >18 | | |
| | Yes n/ % | OR[c] | 95% CI | Yes n/ % | OR | 95% CI | Yes n/ % | OR | 95% CI | Yes n/ % | OR | 95% CI | Yes n/ % | OR | 95% CI | Yes n/ % | OR | 95% CI |
| Untested | 6898/ 3.5 | 0.31 | 0.28-0.34 | 495/ 1.4 | 0.65 | 0.56-0.74 | 1018/ 1.30 | 0.69 | 0.61-0.79 | 564/ 1.0 | 0.39 | 0.34-0.45 | 15/ 1.1 | 0.65 | 0.37-1.13 | 258/ 1.4 | 0.52 | 0.43-0.65 |
| Negative | 929/ 5.8 | 1 | Ref | 575/ 2.4 | 1 | Ref | 398/ 2.0 | 1 | Ref | 459/ 2.2 | 1 | Ref | 704/ 2.4 | 1 | Ref | 234/ 3.4 | 1 | Ref |
| Positive | 2004/ 33.8 | 20.27 | 17.93-22.91 | 587/ 11.4 | 9.65 | 8.31-11.21 | 310/ 7.7 | 6.29 | 5.22-7.57 | 248/ 12.6 | 9.23 | 7.48-11.4 | 80/ 10.8 | 4.86 | 3.57-6.6 | 13/ 15.5 | 4.03 | 1.85-8.8 |
| **Dyspnea (n=530 200, 120 605 participants)** | | | | | | | | | | | | | | | | | | |
| Time since test (months)# | | | | | | | | | | | | | | | | | | |
| **SARS-CoV-2 status** | 0-1 | | | >1-3 | | | >3-6 | | | >6-12 | | | >12-18 | | | >18 | | |
| | Yes n/ % | OR | 95% CI | Yes n/ % | OR | 95% CI | Yes n/ % | OR | 95% CI | Yes n/ % | OR | 95% CI | Yes n/ % | OR | 95% CI | Yes n/ % | OR | 95% CI |
| Untested | 17386/ 8.7 | 0.32 | 0.3-0.34 | 1445/ 4.2 | 0.58 | 0.53-0.64 | 2672/ 3.5 | 0.48 | 0.44-0.52 | 2152/ 3.3 | 0.51 | 0.46-0.55 | 91/ 6.8 | 1.64 | 1.23-2.16 | 664/ 3.5 | 0.6 | 0.54-0.67 |
| Negative | 2212/ 13.8 | 1 | Ref | 1761/ 7.2 | 1 | Ref | 1348/ 6.9 | 1 | Ref | 1380/ 6.7 | 1 | Ref | 1858/ 6.4 | 1 | Ref | 625/ 9 | 1 | Ref |
| Positive | 1765/ 29.7 | 4.65 | 4.17-5.18 | 769/ 14.9 | 4.07 | 3.6-4.6 | 449/ 11.1 | 2.46 | 2.14-2.84 | 247/ 12.6 | 2.8 | 2.32-3.38 | 82/ 11.1 | 2.04 | 1.51-2.75 | 10/ 11.9 | 2.09 | 0.93-4.73 |

**eTable 3 cont.**

| **Fatigue (n=530 200, 120 605 participants)** | | | | | | | | | | | | | | | | | | |
|---|---|---|---|---|---|---|---|---|---|---|---|---|---|---|---|---|---|---|
| Time since test (months)# | | | | | | | | | | | | | | | | | | |
| **SARS-CoV-2 status** | 0-1 | | | >1-3 | | | >3-6 | | | >6-12 | | | >12-18 | | | >18 | | |
| | Yes n/ % | OR | 95% CI | Yes n/ % | OR | 95% CI | Yes n/ % | OR | 95% CI | Yes n/ % | OR | 95% CI | Yes n/ % | OR | 95% CI | Yes n/ % | OR | 95% CI |
| Untested | 34532/ 17.4 | 0.17 | 0.17-0.18 | 3779/ 11.0 | 0.47 | 0.44-0.49 | 8332/ 10.9 | 0.40 | 0.38-0.42 | 7667/ 11.7 | 0.50 | 0.44-0.53 | 227/ 16.9 | 1.16 | 0.97-1.41 | 1982/ 10.5 | 0.44 | 0.41-0.47 |
| Negative | 5617/ 35.0 | 1 | Ref | 5245/ 21.5 | 1 | Ref | 4306/ 22.0 | 1 | Ref | 4359/ 21.0 | 1 | Ref | 6578/ 22.8 | 1 | Ref | 1953/ 28.0 | 1 | Ref |
| Positive | 4236/ 71.4 | 9.46 | 8.65-10.35 | 1735/ 33.5 | 2.86 | 2.62-3.12 | 1182/ 29.3 | 1.87 | 1.69-2.06 | 647/ 32.9 | 2.17 | 1.89-2.48 | 218/ 29.5 | 1.45 | 1.14-1.79 | 26/ 31.0 | 0.9 | 0.50-1.64 |
| **Worsening of health (n=304 958, 120 605 participants)** | | | | | | | | | | | | | | | | | | |
| Time since test (months)# | | | | | | | | | | | | | | | | | | |
| **SARS-CoV-2 status** | 0-1 | | | >1-3 | | | >3-6 | | | >6-12 | | | >12-18 | | | >18 | | |
| | Yes n/ % | OR | 95% CI | Yes n/ % | OR | 95% CI | Yes n/ % | OR | 95% CI | Yes n/ % | OR | 95% CI | Yes n/ % | OR | 95% CI | Yes n/ % | OR | 95% CI |
| Untested | 27/ 13.3 | 0.60 | 0.36-0.98 | 1625/ 11.9 | 0.49 | 0.45-0.53 | 8070/ 10.6 | 0.40 | 0.37-0.42 | 8652/ 13.3 | 0.65 | 0.61-0.68 | 107/ 10.7 | 0.52 | 0.4-0.66 | 2137/ 11.5 | 0.54 | 0.5-0.58 |
| Negative | 2308/ 16.5 | 1 | Ref | 4461/ 18.7 | 1 | Ref | 3740/ 19.4 | 1 | Ref | 3811/ 18.8 | 1 | Ref | 5193/ 18.1 | 1 | Ref | 1432/ 20.7 | 1 | Ref |
| Positive | 1717/ 29.7 | 2.83 | 2.6-3.08 | 1537/ 30.6 | 2.61 | 2.38-2.85 | 1041/ 28.0 | 2.19 | 1.98-2.43 | 618/ 37.1 | 3.56 | 3.09-4.11 | 238/ 36.2 | 3.44 | 2.77-4.26 | 30/ 40.0 | 3.73 | 2.03-6.88 |

**eTable 3 cont.**

| **Fever (n=530 200, 120 605 participants)** | | | | | | | | | | | | | | | | | | |
|---|---|---|---|---|---|---|---|---|---|---|---|---|---|---|---|---|---|---|
| Time since test (months)# | | | | | | | | | | | | | | | | | | |
| **SARS-CoV-2 status** | 0-1 | | | >1-3 | | | >3-6 | | | >6-12 | | | >12-18 | | | >18 | | |
| | Yes n/ % | OR | 95% CI | Yes n/ % | OR | 95% CI | Yes n/ % | OR | 95% CI | Yes n/ % | OR | 95% CI | Yes n/ % | OR | 95% CI | Yes n/ % | OR | 95% CI |
| Untested | 12595/ 6.3 | 0.11 | 0.10-0.12 | 488/ 1.4 | 0.31 | 0.28-0.35 | 1133/ 1.5 | 0.28 | 0.25-0.31 | 890/ 1.4 | 0.22 | 0.19-0.23 | 33/ 2.5 | 0.40 | 0.28-0.58 | 611/ 3.3 | 0.44 | 0.40-0.49 |
| Negative | 2655/ 16.5 | 1 | Ref | 1152/ 4.7 | 1 | Ref | 995/ 5.1 | 1 | Ref | 1247/ 6.0 | 1 | Ref | 2099/ 7.3 | 1 | Ref | 628/ 9.0 | 1 | Ref |
| Positive | 2403/ 40.5 | 4.7 | 4.25-5.18 | 261/ 5.0 | 1.17 | 1.01-1.37 | 210/ 5.2 | 1.06 | 0.9-1.26 | 116/ 5.9 | 0.94 | 0.76-1.17 | 39/ 5.3 | 0.61 | 0.42-0.87 | 5/ 6.0 | 0.54 | 0.20-1.45 |

| **Headache (n=530 200, 120 605 participants)** | | | | | | | | | | | | | | | | | | |
|---|---|---|---|---|---|---|---|---|---|---|---|---|---|---|---|---|---|---|
| Time since test (months)# | | | | | | | | | | | | | | | | | | |
| **SARS-CoV-2 status** | 0-1 | | | >1-3 | | | >3-6 | | | >6-12 | | | >12-18 | | | >18 | | |
| | Yes n/ % | OR | 95% CI | Yes n/ % | OR | 95% CI | Yes n/ % | OR | 95% CI | Yes n/ % | OR | 95% CI | Yes n/ % | OR | 95% CI | Yes n/ % | OR | 95% CI |
| Untested | 52632/ 26.5 | 0.29 | 0.27-0.30 | 5550/ 16.2 | 0.47 | 0.40-0.50 | 13563/ 17.7 | 0.41 | 0.34-0.43 | 14100/ 21.5 | 0.73 | 0.70-0.76 | 274/ 20.4 | 0.95 | 0.80-1.14 | 2887/ 15.4 | 0.43 | 0.41-0.46 |
| Negative | 6566/ 40.9 | 1 | Ref | 7364/ 30.2 | 1 | Ref | 6173/ 31.5 | 1 | Ref | 6026/ 29.1 | 1 | Ref | 9560/ 33.1 | 1 | Ref | 2578/ 36.9 | 1 | Ref |
| Positive | 3703/ 62.4 | 3.34 | 3.08-3.61 | 1311/ 25.3 | 0.8 | 0.73-0.88 | 1100/ 27.2 | 0.73 | 0.66-0.81 | 590/ 30.0 | 0.9 | 0.79-1.03 | 207/ 28.1 | 0.73 | 0.59-0.90 | 32/ 38.1 | 0.93 | 0.54-1.64 |

**eTable 3 cont.**

| **Cough (n=530 200, 120 605 participants)** | | | | | | | | | | | | | | | | | | |
|---|---|---|---|---|---|---|---|---|---|---|---|---|---|---|---|---|---|---|
| Time since test (months)# | | | | | | | | | | | | | | | | | | |
| **SARS-CoV-2 status** | 0-1 | | | >1-3 | | | >3-6 | | | >6-12 | | | >12-18 | | | >18 | | |
| | Yes n/ % | OR | 95% CI | Yes n/ % | OR | 95% CI | Yes n/ % | OR | 95% CI | Yes n/ % | OR | 95% CI | Yes n/ % | OR | 95% CI | Yes n/ % | OR | 95% CI |
| Untested | 37796/ 19.0 | 0.16 | 0.16-0.17 | 3007/ 8.8 | 0.38 | 0.36-0.41 | 6841/ 8.9 | 0.37 | 0.36-0.39 | 5817/ 8.9 | 0.33 | 0.31-0.34 | 127/ 9.4 | 0.35 | 0.28-0.44 | 2506/ 13.3 | 0.53 | 0.50-0.57 |
| Negative | 5985/ 37.2 | 1 | Ref | 4705/ 19.3 | 1 | Ref | 3689/ 18.8 | 1 | Ref | 4305/ 20.8 | 1 | Ref | 7011/ 24.2 | 1 | Ref | 1866/ 26.7 | 1 | Ref |
| Positive | 4040/ 68.1 | 6.05 | 5.57-6.58 | 1136/ 22.0 | 1.45 | 1.32-1.59 | 661/ 16.4 | 0.88 | 0.79-0.98 | 397/ 20.2 | 0.98 | 0.85-1.13 | 140/ 19.0 | 0.70 | 0.55-0.87 | 10/ 11.9 | 0.40 | 0.18-0.85 |

| **Muscular pain (n=530 200, 120 605 participants)** | | | | | | | | | | | | | | | | | | |
|---|---|---|---|---|---|---|---|---|---|---|---|---|---|---|---|---|---|---|
| Time since test (months)# | | | | | | | | | | | | | | | | | | |
| **SARS-CoV-2 status** | 0-1 | | | >1-3 | | | >3-6 | | | >6-12 | | | >12-18 | | | >18 | | |
| | Yes n/ % | OR | 95% CI | Yes n/ % | OR | 95% CI | Yes n/ % | OR | 95% CI | Yes n/ % | OR | 95% CI | Yes n/ % | OR | 95% CI | Yes n/ % | OR | 95% CI |
| Untested | 21711/ 10.9 | 0.30 | 0.28-0.32 | 2643/ 7.7 | 0.56 | 0.50-0.60 | 5587/ 7.3 | 0.48 | 0.45-0.50 | 6059/ 9.3 | 0.70 | 0.67-0.75 | 265/ 19.7 | 2.79 | 2.23-3.39 | 1497/ 8.0 | 0.49 | 0.45-0.53 |
| Negative | 2924/ 18.2 | 1 | Ref | 3083/ 12.6 | 1 | Ref | 2574/ 13.1 | 1 | Ref | 2699/ 13.0 | 1 | Ref | 3949/ 13.7 | 1 | Ref | 1176/ 16.8 | 1 | Ref |
| Positive | 2476/ 41.7 | 6.61 | 6.01-7.27 | 674/ 13.0 | 1.19 | 1.06-1.34 | 499/ 12.4 | 1.1 | 0.96-1.25 | 307/ 15.6 | 1.34 | 1.13-1.59 | 115/ 15.6 | 1.19 | 0.92-1.55 | 17/ 20.2 | 1.09 | 0.55-2.15 |

**eTable 3 cont.**

| **Nasal symptoms (n=530 200, 120 605 participants)** | | | | | | | | | | | | | | | | | | |
|---|---|---|---|---|---|---|---|---|---|---|---|---|---|---|---|---|---|---|
| Time since test (months)# | | | | | | | | | | | | | | | | | | |
| **SARS-CoV-2 status** | 0-1 | | | >1-3 | | | >3-6 | | | >6-12 | | | >12-18 | | | >18 | | |
| | Yes n/ % | OR | 95% CI | Yes n/ % | OR | 95% CI | Yes n/ % | OR | 95% CI | Yes n/ % | OR | 95% CI | Yes n/ % | OR | 95% CI | Yes n/ % | OR | 95% CI |
| Untested | 47174/ 23.7 | 0.21 | 0.2-0.22 | 5372/ 15.7 | 0.52 | 0.49-0.55 | 13401/ 17.5 | 0.53 | 0.51-0.55 | 10788/ 16.5 | 0.53 | 0.5-0.55 | 183/ 13.6 | 0.46 | 0.38-0.55 | 3226/ 17.2 | 0.61 | 0.57-0.64 |
| Negative | 7221/ 44.9 | 1 | Ref | 6735/ 27.6 | 1 | Ref | 5396/ 27.5 | 1 | Ref | 5512/ 26.6 | 1 | Ref | 9194/ 31.8 | 1 | Ref | 2248/ 32.2 | 1 | Ref |
| Positive | 3632/ 61.2 | 2.37 | 2.2-2.56 | 1021/ 19.7 | 0.63 | 0.58-0.69 | 798/ 19.8 | 0.55 | 0.5-0.61 | 460/ 23.4 | 0.75 | 0.65-0.86 | 160/ 21.7 | 0.5 | 0.41-0.63 | 19/ 22.6 | 0.5 | 0.27-0.93 |
| **Sore throat (n=530 200, 120 605 participants)** | | | | | | | | | | | | | | | | | | |
| Time since test (months)# | | | | | | | | | | | | | | | | | | |
| **SARS-CoV-2 status** | 0-1 | | | >1-3 | | | >3-6 | | | >6-12 | | | >12-18 | | | >18 | | |
| | Yes n/ % | OR | 95% CI | Yes n/ % | OR | 95% CI | Yes n/ % | OR | 95% CI | Yes n/ % | OR | 95% CI | Yes n/ % | OR | 95% CI | Yes n/ % | OR | 95% CI |
| Untested | 43638/ 21.9 | 0.13 | 0.12-0.14 | 3696/ 10.8 | 0.44 | 0.42-0.47 | 9594/ 12.5 | 0.48 | 0.46-0.49 | 7328/ 11.2 | 0.47 | 0.44-0.49 | 105/ 7.8 | 0.35 | 0.27-0.44 | 2097/ 11.2 | 0.49 | 0.46-0.52 |
| Negative | 7499/ 46.7 | 1 | Ref | 5642/ 23.1 | 1 | Ref | 4533/ 23.1 | 1 | Ref | 4510/ 21.8 | 1 | Ref | 7552/ 26.1 | 1 | Ref | 1967/ 28.2 | 1 | Ref |
| Positive | 3203/ 54.0 | 1.39 | 1.29-1.50 | 761/ 14.7 | 0.56 | 0.51-0.62 | 635/ 15.7 | 0.55 | 0.50-0.61 | 379/ 19.3 | 0.79 | 0.69-0.92 | 140/ 19.0 | 0.64 | 0.51-0.80 | 16/ 19.1 | 0.71 | 0.37-1.36 |

**eTable 3 cont.**

| Abdominal pain (n=530 200, 120 605 participants) | | | | | | | | | | | | | | | | | | |
|---|---|---|---|---|---|---|---|---|---|---|---|---|---|---|---|---|---|---|
| Time since test (months)# | | | | | | | | | | | | | | | | | | |
| **SARS-CoV-2 status** | 0-1 | | | >1-3 | | | >3-6 | | | >6-12 | | | >12-18 | | | >18 | | |
| | Yes n/ % | OR | 95% CI | Yes n/ % | OR | 95% CI | Yes n/ % | OR | 95% CI | Yes n/ % | OR | 95% CI | Yes n/ % | OR | 95% CI | Yes n/ % | OR | 95% CI |
| Untested | 23707/ 11.9 | 0.52 | 0.48-0.54 | 2693/ 7.9 | 0.71 | 0.67-0.76 | 6250/ 8.2 | 0.69 | 0.65-0.72 | 5035/ 7.7 | 0.70 | 0.66-0.74 | 127/ 9.4 | 1.14 | 0.91-1.43 | 1171/ 6.2 | 0.53 | 0.50-0.58 |
| Negative | 2479/ 15.4 | 1 | Ref | 2944/ 12.1 | 1 | Ref | 2402/ 12.3 | 1 | Ref | 2405/ 11.6 | 1 | Ref | 3622/ 12.5 | 1 | Ref | 1082/ 15.5 | 1 | Ref |
| Positive | 1319/ 22.2 | 1.82 | 1.66-2.0 | 416/ 8.0 | 0.63 | 0.56-0.72 | 377/ 9.3 | 0.71 | 0.62-0.81 | 192/ 9.8 | 0.68 | 0.56-0.82 | 80/ 10.8 | 0.73 | 0.54-0.96 | 10/ 11.9 | 0.50 | 0.22-1.11 |

| Asymptomatic (n=530 200, 120 605 participants) | | | | | | | | | | | | | | | | | | |
|---|---|---|---|---|---|---|---|---|---|---|---|---|---|---|---|---|---|---|
| Time since test (months)# | | | | | | | | | | | | | | | | | | |
| **SARS-CoV-2 status** | 0-1 | | | >1-3 | | | >3-6 | | | >6-12 | | | >12-18 | | | >18 | | |
| | Yes n/ % | OR | 95% CI | Yes n/ % | OR | 95% CI | Yes n/ % | OR | 95% CI | Yes n/ % | OR | 95% CI | Yes n/ % | OR | 95% CI | Yes n/ % | OR | 95% CI |
| Untested | 84733/ 42.6 | 6.37 | 6.15-6.61 | 19835/ 57.8 | 2.33 | 2.23-2.43 | 43031/ 56.2 | 2.36 | 2.27-2.45 | 34403/ 52.5 | 1.58 | 1.52-1.64 | 614/ 45.6 | 0.76 | 0.66-0.88 | 11097/ 59.0 | 2.0 | 1.91-2.1 |
| Negative | 3138/ 19.5 | 1 | Ref | 8945/ 36.7 | 1 | Ref | 7387/ 37.7 | 1 | Ref | 8313/ 40.0 | 1 | Ref | 10301/ 35.6 | 1 | Ref | 2263/ 32.4 | 1 | Ref |
| Positive | 225/ 3.8 | 0.11 | 0.11-0.13 | 1704/ 32.9 | 0.66 | 0.61-0.72 | 1455/ 36.0 | 0.88 | 0.80-0.96 | 533/ 27.1 | 0.51 | 0.45-0.58 | 227/ 30.8 | 0.78 | 0.63-0.95 | 25/ 29.8 | 0.94 | 0.52-1.69 |

* Adjusted for age (10-years categories), gender (men, women) body mass index (<25 kg/ $m^2$, ≥25 kg/ $m^2$), income level per household (< 299 999, 300 000-599 999, 600 000-1000 000, >1000 000 NOK, missing), smoking status (never, former, current, missing), underlying medical condition (no, yes, missing) and symptom status at baseline (no, yes, missing).
### For the untested, time since baseline was used in place of time since a positive- or negative SARS-CoV-2 test.

##### Supplementary eTable 4. The association between the number of days bedridden with COVID-19 reported in the last follow-up questionnaire and each symptom reported at the last follow-up questionnaire.

| Symptom | Bedridden during COVID-19 (days) | Adjusted odds ratio | 95% confidence interval | N |
|---|---|---|---|---|
| Memory problems | 0 | 1 (Ref) | - | 14584 |
| | 0-6 | 2.34 | 2.04 - 2.68 | |
| | 7-13 | 5.34 | 4.29 - 6.64 | |
| | >=14 | 9.12 | 6.51 - 12.77 | |
| Concentration problems | 0 | 1 (Ref) | - | 14584 |
| | 0-6 | 2.15 | 1.94 - 2.38 | |
| | 7-13 | 4.65 | 3.86 - 5.6 | |
| | >=14 | 6.99 | 5.11 - 9.56 | |
| Anosmia or dysgeusia | 0 | 1 (Ref) | - | 14584 |
| | 0-6 | 1.32 | 1.21 - 1.45 | |
| | 7-13 | 1.84 | 1.52 - 2.22 | |
| | >=14 | 1.97 | 1.41 - 2.76 | |
| Dyspnoea | 0 | 1 (Ref) | - | 14584 |
| | 0-6 | 2.07 | 1.88 - 2.27 | |
| | 7-13 | 3.44 | 2.87 - 4.13 | |
| | >=14 | 3.53 | 2.58 - 4.83 | |
| Fatigue | 0 | 1 (Ref) | - | 14584 |
| | 0-6 | 2.09 | 1.94 - 2.24 | |
| | 7-13 | 3.18 | 2.66 - 3.79 | |
| | >=14 | 3.41 | 2.47 - 4.70 | |
| Worsening of self-assessed overall health | 0 | 1 (Ref) | - | 14536 |
| | 0-6 | 2.36 | 2.18 - 2.55 | |
| | 7-13 | 6.07 | 5.1 - 7.21 | |
| | >=14 | 9.42 | 6.79 - 13.08 | |
| Fever | 0 | 1 (Ref) | - | 14578 |
| | 0-6 | 2.26 | 2.06 - 2.49 | |
| | 7-13 | 2.27 | 1.86 - 2.78 | |
| | >=14 | 1.82 | 1.26 - 2.64 | |
| Headache | 0 | 1 (Ref) | - | 14584 |
| | 0-6 | 1.55 | 1.44 - 1.67 | |
| | 7-13 | 1.91 | 1.61 - 2.26 | |
| | >=14 | 1.39 | 1.02 - 1.89 | |

| | | | | |
|---|---|---|---|---|
| Cough | 0 | 1 (Ref) | - | 14584 |
| | 0-6 | 1.27 | 1.18 - 1.36 | |
| | 7-13 | 1.28 | 1.08 - 1.52 | |
| | >=14 | 0.92 | 0.67 - 1.25 | |
| Muscular pain | 0 | 1 (Ref) | - | 14584 |
| | 0-6 | 1.89 | 1.74 - 2.06 | |
| | 7-13 | 2.26 | 1.88 - 2.71 | |
| | >=14 | 1.97 | 1.42 - 2.72 | |
| Nasal symptoms | 0 | 1 (Ref) | - | 14584 |
| | 0-6 | 1.08 | 1 - 1.16 | |
| | 7-13 | 1.04 | .87 - 1.23 | |
| | >=14 | 0.67 | 0.48 -0.93 | |
| Sore troath | 0 | 1 (Ref) | - | 14584 |
| | 0-6 | 10959 | 1.2 - 1.4 | |
| | 7-13 | 1.11 | 0.93 - 1.33 | |
| | >=14 | 0.82 | 0.58 - 1.16 | |
| Abdominal pain | 0 | 1 (Ref) | - | 14578 |
| | 0-6 | 1.73 | 1.55 - 1.93 | |
| | 7-13 | 2.78 | 2.26 - 3.41 | |
| | >=14 | 1.73 | 1.16 - 2.59 | |
| Asymptomatic | 0 | 1 (Ref) | - | 14578 |
| | 0-6 | 0.67 | 0.61 - 0.72 | |
| | 7-13 | 0.43 | 0.33 - 0.54 | |
| | >=14 | 0.57 | 0.38 - 0.85 | |

*The results of a logistic regression model with the symptom as dependent variable and the number of days bedridden as a categorical independent variable. Adjusted for age (10-years categories), gender (men, women) body mass index (<25 kg/ $m^2$, ≥25 kg/ $m^2$), income level per household (< 299 999, 300 000-599 999, 600 000-1000 000, >1000 000 NOK, missing), smoking status (never, former, current, missing), underlying medical condition (no, yes, missing) and symptom status at baseline (no, yes, missing).

#### Supplementary eMethods 1. Substudy between follow-up three and four.

Between the third and fourth follow-up we conducted, an extra follow-up (termed "substudy") that included all participants who had tested positive at that time, as well as a randomly selected group of those who tested negative or were untested. This sub-study included all questionnaire items from the other follow-ups and additionally incorporated both the BRIEF-A and EMQ instruments, which added 88 additional questions.

The objective of this substudy was twofold: to obtain an additional measurement point of symptoms and data on BRIEF-A and EMQ, and to assess the impact of non-response on our overall study by contacting the participants that did not respond after the usual electronic reminders by phone.

To ensure maximum participation, we first sent multiple electronic reminders as for all follow-ups. Thereafter, we released a condensed version of the sub-study questionnaire, excluding the BRIEF-A and EMQ components and finally, we contacted a random subset of the remaining non-responders by phone ("phone cohort").

#### Supplementary eMethods 2. SARS-CoV-2 antibody analysis

A flow cytometer-based method was used to identify IgG antibodies against SARS-CoV-2-derived recombinant antigens in residual sera.[1] Samples with antibodies against either the receptor binding domain (RBD) and full-length spike protein of SARS-CoV-2.[2] A nationwide seroprevalence study in 2021 showed a seroprevalence of 0.9% by January 2021.[3] At the same time, we had registered 936 positive participants, corresponding to a prevalence of 0.8% in our population

1. Holter JC, Pischke SE, de Boer E, et al. Systemic complement activation is associated with respiratory failure in COVID-19 hospitalized patients. *Proc Natl Acad Sci U S A*. Oct 6 2020;117(40):25018-25025. doi:10.1073/ pnas.2010540117
2. Tran TT, Vaage EB, Mehta A, et al. Titers of antibodies against ancestral SARS-CoV-2 correlate with levels of neutralizing antibodies to multiple variants. *NPJ Vaccines*. Dec 30 2022;7(1):174. doi:10.1038/ s41541-022-00586-7
3. Anda EE, Braaten T, Borch KB, et al. Seroprevalence of antibodies against SARS-CoV-2 in the adult population during the pre-vaccination period, Norway, winter 2020/ 21. *Eurosurveillance*. 2022;27(13):2100376. doi:doi:https:/ / doi.org/ 10.2807/ 1560-7917.ES.2022.27.13.2100376

#### Supplementary eMethods 3. Questions from the questionnaires.

**I was tested for COVID-19 with a throat or nose swab** (Complete this question even if you have previously told us that you have had a positive test):

□ Yes, at least one test was positive, and I have/ have had COVID-19
□ Yes, but the test was negative
□ Yes, and I have not received the test result yet
□ No, I have not been tested

**Approximately date when you had your first COVID-19 test?**

(Date field, DD.MM.YYYY, question for those ticking Yes, and at least one test was positive or Yes, I am waiting for the result)

**Check off every symptom you have experienced in the last three weeks.**

**You can tick several symptoms.**

□ Fever
□ High fever (measure to be higher than 39 °C)
□ Heavy breathing (dyspnea)
□ Cough
□ Fatigue
□ Muscle aches
□ Sore throat
□ Impaired sense of smell/ taste
□ Stuffy or runny nose
□ Headache
□ Stomach pain/ nausea/ diarrhoea
□ Memory problems
□ Problems with concentration or thinking
□ Other symptoms
□ No symptoms

**Explain which other symptoms you have experienced the last three weeks:**

(Open text field, question for those ticking other symptoms)
**Compared to a year ago, or before you had COVID-19, how will you describe your health?**

If you have not had COVID-19, think about how it was a year ago.
If you have had COVID-19, think about how it was before you were sick.
□ A lot better now than a year ago
□ A little better now than a year ago
□ About the same as a year ago
□ A little worse than a year ago
□ A lot worse than a year ago

**Sex**

□ Male
□ Female
□ Other

**Enter your age:**

(Open number field)

**What is your height in centimeters?**

(Open number field)

**What is your weight in kilos?**

(Open number field)

**Do you smoke?**

□ Yes
□ No, I have never smoked
□ I have smoked previously
□ I vape
□ Don’t know

**What is the household's annual income (NOK), calculated before taxes?**

□ <299 999 NOK
□ 300 000-599 999 NOK
□ 600 000-1 000 000 NOK
□ >1 000 000 NOK

**Check off any illnesses or conditions you have.**

**You can tick only one option per condition.**

Chronic heart disease including congenital heart disease (not high blood pressure)
□ Yes
□ No
□ Don’t know

High blood pressure
□ Yes
□ No
□ Don’t know

Chronic lung disease (other than asthma)
□ Yes
□ No
□ Don’t know

Asthma
□ Yes
□ No
□ Don’t know

Diabetes
□ Yes
□ No
□ Don’t know

On immunosuppressive treatment
◻ Yes
◻ No
◻ Don’t know

Cancer (being treated)
◻ Yes
◻ No
◻ Don’t know

**Have you been hospitalized due to COVID-19?**

(Question for those ticking Yes, and at least one test was positive, and I have/ have had COVID-19
◻ Yes
◻ No

**Were you bedridden due to your episode of COVID-19?**

◻ No
◻ Yes, 1-6 days
◻ Yes, 7-13 days
◻ Yes, two weeks or more
◻ I do not wish to answer

#### Supplementary eMethods 4. Mixed model regression.

The regression model used to estimate OR and 95%CI in Table 2 is a logistic mixed effect model with random intercept:

$$logit(P(Y_{ij} = 1 \mid b_i)) = \beta_0 + X_{ij}\beta + Z_{ij}\eta + b_i$$

Here $Y_{ij}$ = 1 denotes the presence of a given symptom for subject i (i = 1,...,120 605) when answering questionnaire j (j = 1,...,5). $X_{ij}$ is a combination of test status and time since test at time j (i.e. time of answering questionnaire j) for individual i. $X_{ij}$ is coded as:

1 - untested and 0-1 month since inclusion,
2 - untested and 1-3 months since inclusion,
3 - untested and 3-6 months since inclusion,
4 - untested and 6-12 months since inclusion,
5 - untested and 12-18 months since inclusion,
6 - untested and >18 months since inclusion,
7- negative and 0-1 month since test,
8 - Reference: negative and 1-3 months since the test,
9 - negative and 3-6 months since the test,
10 - negative and 6-12 months since the test,
11 - negative and 12-18 months since the test,
12 - negative and >18 months since the test,
13- positive and 0-1 month since test,
14 - positive and 1-3 months since the test,
15 - positive and 3-6 months since the test,
16 - positive and 6-12 months since the test,
17 - positive and 12-18 months since the test,
18 - positive and >18 months since the test.

Time since test is defined as time between date of answering questionnaire j and date of first positive or negative test (for tested participants, with a positive test superseding a negative test) or time between date of answering baseline questionnaire and date of answering questionnaire j (for untested participants). $Z_{ij}$ is the vector of regression coefficients for adjustment variables with corresponding regression coefficients $\eta$. $exp(\beta)$ is the ORs reported in Table 2 and $b_i$ the subject specific random intercept. In particular the model is given by:

$$\begin{aligned} logit\left(P\left(Y_{ij} = 1 \mid b_i\right)\right) \\ &= \beta_0 + X_{ij}\beta + sex_{ij}\eta_{sex} + BMI_{ij}\eta_{BMI} + income_{ij}\eta_{income} \\ &+ smoking_{ij}\eta_{smoking} \\ &+ underlyingMedicalCondition_{ij}\eta_{underlyingMedicalCondition} \\ &+ baselineSymptomStatus_{ij}\eta_{baselineSymptomStatus} + b_i \end{aligned}$$

All included independent variables are categorical, thus $\beta$ and $\eta_.$ are vectors of regression coefficients. The adjustment variables are coded as follows: age (10-year categories, ref 18-29), gender (men, women, ref men), body mass index (BMI,<25 kg/ m2 >25 kg/ m2, missing, ref <25 kg/ m2), annual household income level (< 299 999, 300 000-599 999, 600 000-100 0000, >1 000 000 NOK, missing, ref <299 999), smoking status (never, former, current, missing, ref never), underlying medical condition (no, yes, missing, ref no) and symptom status for each symptom at baseline (no, yes, missing, ref no) or at the first questionnaire asking about the symptom, but before the outcome symptom. The SF 36 health-transition question on self-assessed worsening of overall health was dichotomized from the original 5-levels.

The data consists of repeated measurements for all participants. Symptoms were “measured” at each answered questionnaire, and test status and time since test were allowed to change over questionnaires. The correlation between measurements from the same participant was accommodated with inclusion of a random intercept in the logistic regression model. The random intercept model assumes heterogeneity in the participants' underlying risk of experiencing a symptom which persists over all answered questionnaires (i.e. the model assumes and accommodates dependence in a symptom within individuals), however given the random intercept the symptom is assumed independent.

#### Supplementary eMethods 5. Moving average of symptom prevalence before, during and after test (Figure 1, eFigure 3, 4 and 5).

To illustrate the average prevalence of symptoms before and after the day of testing and in the period around testing we calculated the moving average symptom prevalence (Figure 1). Unlike in the statistical model, the positive and negative participants do not change status during the study period in this figure. So, in the figure all responses from participants who at some point during the study period have a positive SARS-CoV-2 test are represented in the positive line. Responses from participants who have tested negative and never tested positive are represented in the negative line. Untested participants do not have a specific time of testing. Instead, they are represented by the red-dashed horizontal line, which indicates their average proportion of symptoms across all time-points throughout the entire study period. Note that all questionnaires with the untested status are included in this average - this also includes responses from participants that at a later point in time will become either negative or positive.

To capture the strong fluctuations in symptoms around a positive or negative test, a shorter interval around day 0 (day -50 to 100) was used to prevent excessive smoothing.

To calculate the average prevalence of symptoms before, and after testing, we calculated the average prevalence over 84 days period from day -500 to -50 before testing and from day 100 to 600 after testing. For the testing period (day -50 to 100, we used a shorter 21-day interval.

#### Supplementary eMethods 6. Stratified analyses of the time bedridden

The bedridden question was only given to SARS-CoV-2 positive participants. To calculate the adjusted odds ratios with 95% confidence intervals for each category of bedridden during COVID-19 (0 days (reference), 1-6 days, 7-13 days, or >=14 days), a logistic regression model with each symptom reported in the last follow-up questionnaire as dependent variable, bedridden reported in the last follow-up questionnaire as an independent categorical variable were applied. The same potential confounders as in the mixed model analyses were included: age (10-year categories), gender (men, women), body mass index (BMI,<25 kg/ m2 >25kg/ m2, missing), annual household income level (< 299 999, 300 000-599 999, 600 000-100 0000, >1 000 000 NOK, missing), smoking status (never, former, current, missing), underlying medical condition (no, yes, missing) and symptom status for each symptom at baseline (no, yes, missing) or at the first questionnaire asking about the symptom, but before the outcome symptom.
